# Rapid changes in population socioeconomic status indicators are unevenly distributed in a rural Pakistani village

**DOI:** 10.64898/2026.01.22.26344397

**Authors:** Syed Iqbal Azam, Zeba A Rasmussen, Ejaz Hussain, Natalie M. Chen, Wasiat H Shah, Benjamin JJ McCormick, the Oshikhandass Water, Sanitation, Health and Hygiene Interventions Project

## Abstract

**Background:** Socioeconomic status is an important driver of health outcomes, but the drivers of change in household status over time are often overlooked in favor of cross-sectional metrics of convenience. This is especially challenging in remote populations in low– and middle-income settings. We report changes in socioeconomic status over two decades.

**Methods:** During two studies on childhood health outcomes (1989-1996 and 2011-2014), socioeconomic data were collected through surveys and interviews in Oshikhandass village, a remote population in northern Pakistan. Observations included metrics of population demography, occupation and incomes, household structure and self-reported adult illness. Individual measurements are reported with summary statistics comparing the two time periods and multi-dimensional socioeconomic constructs are constructed from factor analyses to compare changes in relative household socioeconomic position over time.

**Results:** The population had substantial investment during the early 1990s that specifically targeted suspected causes of poverty (electrification, clean water and female education). Within one generation, the village approximately doubled in population, was fully electrified, maternal illiteracy rates dropped from 70% to 27% and jobs opportunities proliferated from primarily agricultural to include a large number of service sectors. Simultaneously the population demography has transformed from stage 1 to stage 3 unlike the national trend that remains in stage 2. Despite secular trends to improved status, there has been approximately equal decline in relative position in the population due to small changes in access to resources.

**Conclusion:** The transformation in human capital in this population is a testament to targeted investments to improve education and childhood health. In a single generation the population has transitioned faster than the national average, exemplified, for example, by individuals achieving higher education, including international, and the village becoming a magnet for migration from the surrounding population. The transition has not been evenly distributed, however, and access to land and resources have led to some households rising in relative position while others have fallen.

## 1 Introduction

Socioeconomic status (SES) describes an individual’s ability to access economic and social resources within a population. SES is a dominant driver of health outcomes, and there is a long history evidencing how individuals characterized by lower SES tend to have poorer health outcomes (Winkleby et al. 1992, Adler et al. 1993, House, Kessler, and Herzog 1990, Gupta et al., 2017). However, despite overwhelming evidence that SES is important, there is considerable variability in how it is measured. Selecting appropriate SES indicators is challenging both because it is a multifaceted construct and because socioeconomic relationships and resources vary over time (Duncan et al. 2002, Durkin, 1994).

An individual’s SES is a complex function of multiple dimensions that can change at different rates and extents; for example, access to resources may be determined by relatively static factors such as gender, slowly changing factors like household location or structure, and factors that can change rapidly at some points and slowly at others such as education, occupation, and income. Indicators are variably used individually or in statistical combinations, although the link between SES and health is most established using observations of educational attainment (e.g., Winkleby et al. 1992, Howe et al. 2012), income (e.g., Duncan et al. 2002), and occupation (e.g., Fujishiro et al, 2010).

Globally, Pakistan ranks 168 out of 193 countries in the 2025 Human Development Index (UNHDR HDI, 2025). This index captures key indicators of SES (e.g. education, income) and combines them with health metrics (e.g. child mortality). Relative to global development, Pakistan has performed poorly despite steady improvement: child mortality rates and undernutrition (Globalhungerindex.org/Pakistan.html, 2025) have remained high while life expectancy, educational attainment and adult literacy have remained low relative to other countries. However, within the country, there is considerable heterogeneity, not least because of substantial wealth and income inequalities, especially for females (Khan, 2020, World Inequality Database, 2025). Within any one community there are opportunities for better or worse health outcomes as people stratify along a continuum of SES. The northeastern mountains of Pakistan have a population that has undergone dramatic social changes, especially in educational opportunities (Benz, 2013, Rasmussen et al., 2021).

This study investigates changes in household-level SES, including education and occupation over a 20-year period and aims to contextualize changes in SES and health in a village in the Gilgit-Baltistan (G-B) region of Pakistan. This area is physically remote despite being in close proximity to international borders for Afghanistan, China, and India. After the opening of the Karakoram Highway in 1978, connecting this area to Pakistan’s capital and the south, as well as north to China, this strategically important area with a historically low base in many aspects of SES has seen major investments in health and other development expenditures by both the Government of Pakistan and civil society organizations, including those of the Aga Khan Development Network (WB report, 2010, IBRD, 1990). This study leverages detailed data collection two decades apart to paint a portrait of the transition in SES in a village community near Gilgit, the administrative capital of G-B. We do this to help contextualize changes in health outcomes which have been described in other studies including dramatically decreased infant and child mortality and increased life expectancy (Walraven 2009, Hansen et al., 2020).We hypothesize that households of long standing in the community have had greater increases in dimensions of SES around opportunities and social status, but that their SES around their physical household environment is no better than households that have migrated into the population, which benefit from secular trends in, for example, community infrastructure and construction techniques and technology. We discuss the implications of the SES transition in this population as a driver for future health care needs.

## 2 Methods

There were two study periods, the first from 1989-1996 (hereafter, Study 1) and the second from 2011-2014 (hereafter, Study 2), described elsewhere (Hansen et al., 2020, Rasmussen et al., 2021). Study 1 focused on surveillance of child health and had few variables related to SES, informed by knowledge at the time, but included information on house construction, and water and sanitation facilities. Study 2 was an opportunity to revisit the population, this time again focusing on childhood diarrhea and pneumonia, and follow up of children from Study 1 as adolescents and young adults. After nearly two decades there was a greater focus on SES, and variables included levels of education, utilization of water and fuel, and household assets including livestock and land.

### 2.1 Study Area

The village of Oshikhandass is roughly 20 km from Gilgit, the region’s economic hub. The village’s health services are delivered at the local government dispensary or at hospitals in Gilgit (Hussein and Khan, 2010). Historically, the local economy operated through small-scale agriculture (AKRSP 1987), but the implementation of national and regional programs in education, water and sanitation infrastructure (Ahmed et al., 1996), (AKRSP 1990, 1995), and social and economic mobilization (AKRSP 1987) resulted in substantial social and economic development over the study period in the village population (Rasmussen et al., 2021). In 1994, hydro-generated electricity arrived in the village, but was irregular during winter months when water sources were frozen. The main languages in the village were Shina, Brusheski, and Urdu (the national language), which was spoken by educated individuals.

### 2.2 Household Data

In July 1989, all structures in the village were mapped, and study staff and community research workers conducted a door-to-door census to determine the number of individuals in each structure, defined as those sharing a kitchen. Multiple families could live in the same structure; a family/household was defined as a mother, father and children, along with grandparents and other related individuals who were living together. This census was updated every six months from 1990-1996. Data were collected on age of individuals, type of household construction (unimproved or mud/stone; partially improved or mud/stone with some cement; improved or cement), number of rooms (excluding kitchen, veranda, storeroom and latrine), type of toilet (unimproved traditional called *chukun* or field; flush), location of water source (on premises or not), treatment of water (none, yes), type of lighting (candle; kerosene lantern; gas), and type of fuel used for heating/cooking (e.g., wood, kerosene or gas).

In July 2011, another village census was conducted, identifying participants from the original study who still lived in Oshikhandass and those who had migrated into the village since 1996. Interviews were conducted in Urdu or in the participant’s mother tongue by study staff in person in the participant’s home.

A questionnaire (see Supplementary Appendix 1) was used to collect data on socioeconomic status, including additional detailed information that was not envisaged in Study 1. All questionnaires were translated into Urdu. Data were collected on: highest level of education in the household; type of house construction (material used in roof, walls); number of total rooms and number used for sleeping; whether the household had electricity; alternative sources of power/lighting; types of fuel for heating; type of cooking stove and fuel used for cooking; type of ventilation; type of toilet (traditional *chukun* or field, pit latrine; improved flush to septic tank or pit latrine); access to water from filtration plant (none, access to non-functional plant, or to functional plant); water delivery (piped to dwelling or not); source and location of drinking water; treatment and storage of water. Some households in Study 2 were only present during the initial mapping and were asked a more limited number of questions on water and sanitation.

### 2.3 Demographics and health

In both studies, data on births and deaths of children under age five years were collected weekly. In Study 1, deaths of other household members were collected every six months, but systematic supervision was not conducted as adult deaths were not the focus of the study. In Study 2, data were collected on individuals who had died in the year before registration, with reported cause of death.

In both studies, data were collected on diarrhea and pneumonia in children under age five (Hansen et al., 2020). In Study 2, data were collected through a questionnaire (see Supplementary Appendix 2) of household family members with cancer, diabetes, hypertension, cardiac diseases, blood disorders, genetic disease, mental disorders, and birth defects; other conditions were coded later. Data were collected on acute and chronic medical conditions and cost of treatment in the last month, health-related decision making, and smoking in the home.

In Study 1, data were collected on those migrating into and out of the village. In Study 2, data were collected on those who had migrated into and out of the village since Study 1 ended in 1996. We did not collect data on religious background and were therefore unable to explore these cultural influences on SES.

### 2.4 Occupation and Education

Data on occupation and monthly income were collected in both studies from parents of children under age 5 years (see Supplementary Appendix 3). In Study 2, additional data were collected on all earning adults in households including type and location of employment. Additional questions in Study 2 covered: household assets; ownership and of number of animals, home and amount of agricultural and non-agricultural land; monthly income and savings; and financial decision-making.

### 2.5 Statistical Analysis

R (version 4.5.1) and Stata (version 15.0) were used to analyze the data.

#### 2.5.1 Demographics

Census data from the two study periods were used to construct population pyramids following Demographic and Health Survey Methods (Pakistan Demographic and Health Survey 2012-13, 2013).

#### 2.5.2 Health Outcomes

The frequency of health conditions reported in Study 2 by those over 18 years of age who were still living and for those who were reported to have died in the previous year were determined. The difference between health conditions and sex was tested by chi square.

#### 2.5.2 Occupation and Education

Descriptive statistics were calculated for the frequency of different occupations and to compare the composition of households in the two study periods. Occupations were listed according to the Pakistan Census Standard Classification of Occupations (1981-1998); these were categorized into high, medium and low occupational status by local experts based on perceived status.

#### 2.5.3 SES Scores

Three SES scores were created (Figure 1) using factor analyses to create latent SES variables (i.e., constructs that are not directly measurable) from underlying patterns in the multivariate data. The first two SES scores used only those variables common to both Study 1 and Study 2, starting with an Exploratory Factor Analysis of the data from Study 1 data, then applying the same factor structure in a Confirmatory Factor Analysis to data from Study 2 to detect changes in SES; the third score used all the additional information collected only in Study 2 to generate a more detailed description of SES.

**Figure 1:**
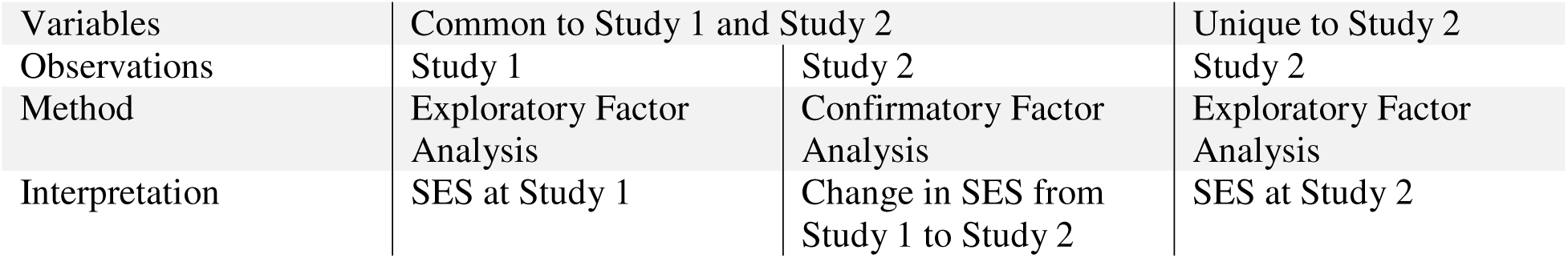
Socioeconomic status (SES) variables for each study timepoint. The first two metrics allow comparison of change over time because they are based on the same variables. The third metric includes more variables for a richer description of SES.

The variables common to Study 1 and 2 were: dwelling construction, lighting source, primary source of cooking fuel, number of rooms (total rooms in Study 1, used for sleeping in Study 2), toilet facility and piped water supply on premises (Study 1) or piped to household (Study 2). Additional variables used for analysis in Study 2 included those mentioned in Section 2.4 and listed in Supplementary Table 1.

The steps taken to create composite SES metrics were as follows: ordinal categorical variables were manually coded, using expert opinion, to give an ordered rank. For example, drinking water available inside the home was ranked higher than water that had to be brought in; similarly, concrete construction was ranked higher than a wooden structure. Ranking reflected perceived wealth status, *i.e.*, the higher the ordinal coding, the higher the SES. Nominal categorical variables were separated into binary yes/no variables.

Factor analyses were used to calculate a simplified multidimensional score for SES. The Kaiser-Meyer-Olkin measure of sampling adequacy and Bartlett’s test of sphericity were used to assess the appropriateness of the data for factor analysis. Exploratory Factor Analysis (EFA) was used to obtain factor loadings by using principal component factor and iterated component factor without rotation and then with orthogonal varimax (Kaiser off) rotation. Maximum likelihood extraction and Eigenvalues ≥1 were used to retain factors. Finally, only variables with loadings ≥0.3 were kept to create an aggregate SES score variable. Confirmatory Factor Analysis (CFA) was used to apply the EFA from Study 1 to the data of Study 2. An additional SES score using CFA of all the variables in Study 2 was calculated. The three aggregate SES scores were divided into quintiles (from lowest to highest).

Individual elements of SES were compared to the household SES percentile using analysis of variance (ANOVA) and where statistically significant (assuming p < 0.05).

### 2.6 Ethical approval

Ethical approval was granted for Study 1 by the Aga Khan University Human Subjects Protection Committee (November 15, 1989). Parents/legal guardians of children provided oral informed consent to participate, given high levels of illiteracy. Ethical approval for Study 2 was granted by the US National Institutes of Health Institutional Review Board (#20TWN071, initially approved by the National Institute of Child Health and Development’s institutional review board), the Aga Khan University Ethics Review Committee (1966-CHS-ERC-11), and the Karakoram International University Ethics Review Committee. All participants provided signed consent to participate.

## 3 Results

### 3.1 Household changes

Table 1 gives characteristics of the full sample of 807 households in Study 1 and 973 households in Study 2. There were n=652 households who were present in both study periods and were used in the analyses to compare the two time-points. The number of households increased by approximately 20% between the two timepoints, mainly due to migration into the village between 1996 and 2011. There were substantial improvements in the building construction (mud/stone to cement/brick), toilet (traditional to flush toilets), electricity access, and access to filtered and piped water. Households were on average one room larger in Study 2 compared to Study 1, had fewer individuals living per structure and per family, with lower occupancy density (individuals per structure per room) reflecting that there were more families per structure in Study 1. However, cooking fuel remained similar (mostly wood). Water treatment was higher in Study 1, but this was primarily by letting glacier water settle silt, while in Study 2, almost all treatment was by boiling.

**Table 1:** Study 1 and 2 household characteristics, water & sanitation.

|  | Study 1 |  | Study 2 |
| --- | --- | --- | --- |
| **n = 807 Households (HH)** |  | **n = 900 Households (HH)<sup>1</sup>** |  |
| ***Dwelling Construction (%)*** |  |  |  |
| Cement | 176 (21.8) | Cement/brick | 474 (51.4) |
| Stone (some cement/mud) | 367 (45.5) | Stone | 438 (47.5) |
| Mud/Stone only | 263 (32.6) | Mud, Wood, No Walls | 11 (1.2) |
| ***Toilet Facility (%)*** |  |  |  |
| Flush | 35 (4.3) | Flush (n=973)<sup>2</sup> | 605 (65.6) |
| Pit Latrine | 102 (12.7) | Pit Latrine | 10 (1.1) |
| *Chukun*<sup>3</sup>/Field | 669 (83.0) | *Chukun*<sup>3</sup>/Field (n=973) | 297 (32.2) |
| Electricity (%) | 0 |  | 917 (99.6) |
| ***Lighting Source (%)*** |  |  |  |
| - |  | Generator | 31 (3.3) |
| Gas | 52 (6.5) | Gas | 251 (27.2) |
| Kerosene | 727 (90.8) | Kerosene | 0 |
| - |  | Rechargeable Lights | 518 (56.2) |
| Candles | 22 (2.7) | Candles, Lantern | 122 (13.2) |
| ***Drinking Water (%)*** |  |  |  |
| Tank | 38 (4.7) | Improved Filtration Plant | 353 (38.6) |
| Storage pit (*gulk*) for water from stream) | 665 (82.5) | Non-Functioning Filtration Plant | 217 (23.7) |
| Stream/River/Spring | 103 (12.8) | Channel/Spring | 345 (37.7) |
| Treat Drinking Water | 442 (54.9) | Treat Drinking Water | 427 (46.3) |
| ***Cooking Fuel (%)*** |  |  |  |
| Gas | 17 (2.1) | Gas (n=973) | 65 (7.1) |
| Electricity | 0 | Electricity (n=973) | 5 (0.5) |
| Kerosene | 17 (2.1) | Kerosene (n=973) | 0 |
| Wood | 772 (95.6) | Wood (n=973) | 851 (92.4) |
| **Household Characteristics (median, IQR)** |  |  |  |
| Persons living per structure, n=463 | 11.00 (8.00, 14.75] |  | 7 (6, 10) |
| Persons living per HH | 7 [4, 9] |  | 6 (5, 7) |
| HH per structure | 1(1,2) |  | 1 [1,2] |
| Maximum Household Size | 8 [7, 10] |  | 6 [5,7] |
| Structure Density<sup>4</sup> | 6.00 [4.00, 9.00] |  | 3.5 (2.5, 5) |
| Household Density<sup>5</sup> | 3.00 [1.67, 5.00] |  | 3 (2, 4) |
| Children <5 years per HH | 3 [2,4] |  | 1 (0, 2] |
| Number of Rooms | 2.00 [1.00, 3.00] |  | 3.00 [2.00, 4.00] |

<sup>1</sup> 900 households completed SES questionnaire; <sup>2</sup> Includes all 973 households with data from SES questionnaire and mapping; <sup>3</sup> Traditional latrine; <sup>4</sup> Persons living per structure/room; <sup>5</sup> Persons living per family/room.

### 3.2 Demographic changes

#### 3.2a. Population Age Distribution

The population grew between Study 1 (n=3669) and Study 2 (n=7570), but in so doing the population has undergone rapid transition (**Figure 2**). The age distribution has shifted from a population with a high birth rate and more than 21% of the population under five years old (over 50% was below 15 years), to a population with lower birth rate and more individuals of working age (only 10.6% were under five and less than a third were under 15 years old). In Study 1, there were more children under age five per household (median 3) than in Study 2 (median 1). In Study 2, there were proportionately more older adults, in particular older females, than in Study 1 although the sex ratio remained similar (1.08 and 1.09 males to females in Study 1 and 2 respectively).

**Figure 2.**
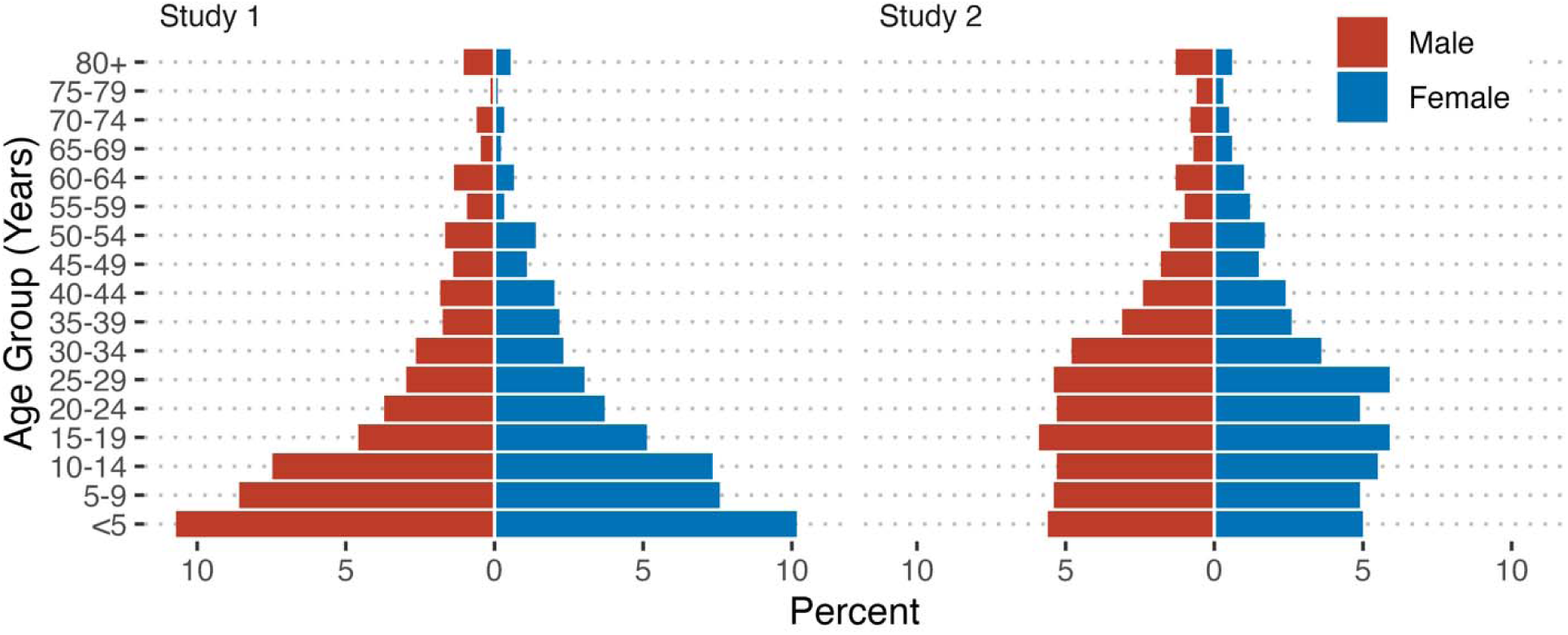
Population pyramids indicating percentage of the study populations stratified by age and sex.

#### 3.2 b Migration

During Study 1, from 1989 to 1996, 749 individuals migrated in and 293 migrated out of Oshikhandass. From 1996 to 2011, 125 households including 619 individuals moved out of the village. During the same period, 186 households with 1506 individuals moved in. Most of these were from nearby and a few from distant areas which were not generally as prosperous as Oshikhandass, including Bagrote (66 households, 16.9% of total), Astore (42, 10.7%), Yaseen (19, 4.9%), Haramosh (15, 3.8%), and Baltistan (12, 3.1%). Most of these households (137) moved in from 2003-2011. The influx of a large number of households contributed to the population increase.

### 3.3 Health Outcomes from Study 2

The distribution of illnesses across adults living with illness and reported deaths is shown in **Table 2**. Fewer than five percent of adults reported living with any of the chronic diseases queried. Hypertension was the most commonly reported condition (3.4% of adults) and the leading reported cause of death (32.3%). Although cardiac disease was more frequently reported by men than women, it was not significantly different (living with illness, p = 0.78; death, p = 0.07). Among those living at the time of the study, females had a statistically significantly higher rate of hypertension (p=0.005).

**Table 2.** Self-reported adults (age >18 years) living with various health conditions (n=4412) or family-reported deaths in the past year in Study 2.

| Type of Illness Reported | Living with illness |  |  | Died |  |  |
| --- | --- | --- | --- | --- | --- | --- |
|  | N(%) |  |  | N(%) |  |  |
|  | Male | Female | Both | Male | Female | Both |
| N | 2343 | 2069 | 4412 | 53 | 43 | 96 |
| Hypertension | 61 (2.6) | 89 (4.3) | 150 (3.4) | 14 (26.4) | 17 (39.5) | 31 (32.3) |
| Cardiac Disease | 8 (0.34) | 4 (0.19) | 12 (0.27) | 19 (35.9) | 7 (16.3) | 26 (27.1) |
| Cancer | 1 (0.04) | 2 (0.10) | 3 (0.07) | 13 (24.5) | 13 (30.2) | 26 (27.1) |
| Diabetes | 13 (0.55) | 14 (0.68) | 27 (0.61) | 5 (9.4) | 4 (9.3) | 9 (9.4) |
| Genetic Disease | 4 (0.17) | 5 (0.24) | 9 (0.20) | 4 (7.6) | 2 (4.7) | 6 (6.3) |
| Mental Disorder | 13 (0.55) | 2 (0.10) | 15 (0.34) | 1 (1.9) | 1 (2.3) | 2 (2.1) |
| Blood Disorder | 2 (0.09) | 4 (0.19) | 6 (0.14) | 1 (1.9) | 0 (0.0) | 1 (1.0) |
| Birth Defects | 7 (0.30) | 6 (0.29) | 13 (0.29) |  |  |  |
| Asthma | 1 (0.04) | 3 (0.14) | 4 (0.09) |  |  |  |
| Tuberculosis | 0 (0.0) | 1 (0.05) | 1 (0.02) |  |  |  |
| Stroke | 1 (0.04) | 0 (0.0) | 1 (0.02) |  |  |  |
| Other Illnesses<sup>1</sup> | 1 (0.04) | 3 (0.14) | 4 (0.09) |  |  |  |

<sup>1</sup> Other Illnesses including malformed legs, paralysis, joint body pain, and body swelling.

Data were available for 900 of 973 households in Study 2; 102/900 (11.3%) reported having an acute or chronic illness in the previous month. The median cost of treatment was 10,000 Pakistani Rupees (PKR, 93.5PKR = 1USD in 2012) (IQR 4000, 50000). Most were able to pay this from savings (74) while the remainder had to take loans from family/friends (16), a bank (9), their village organization (1), or a combination. One household had to sell land to pay a medical bill of PKR 200000. Only in a small number of cases (14/102, 13.7%), illness reduced the working productivity of an earning family member. Usually in medical emergencies, the final decision-maker was the head of household (typically a male), but it could be his wife, daughter/daughter-in-law, son or mother. Usually, vans were used as the main means of transportation, followed by cars. More than a fifth of households, 202/900 (22.4%) reported that household members smoked cigarettes, with usually one member smoking, and most smoking within the house.

### 3.4 Education

Educational status of parents of children under age five was compared for Study 1 and 2 (Table 3). For mothers, there was a major change in educational achievement: the percentage illiterate dropped from nearly 70% to 27%, while the percentage who had done secondary education or above increased from 20% to 65%, including 18% who had completed graduate level education (14 years) or a professional degree. For fathers, there were also shifts to higher educational status. The illiteracy rate dropped from 33% to 11%; 56% were in secondary school or had completed 11 or 12 years of education (Classes 11-12), and 21% had graduated or had a professional degree. Fathers continued to be more educated than mothers in each of the higher categories, with the exception of the percentage who were graduates (mothers 13.4%, fathers 12.9%).

**Table 3.** Education of parents of children under age five – Study 1 and Study 2.

|  | **Study 1** | **Study 2** |
| --- | --- | --- |
| **Mother's Highest Level of Education [n (%)]** | **641<sup>1</sup>** | **677<sup>2</sup>** |
| Illiterate/No formal education | 448 (69.9) | 185(27.3) |
| Primary (up to 5 Years) | 68 (10.6) | 54 (8.0) |
| Secondary/Matric/Class 9-10 | 100 (15.6) | 201 (29.7) |
| Intermediate | 21 (3.3) | 112 (16.5) |
| Graduate (14 years total education) | 2 (0.3) | 91 (13.4) |
| Professional degree<sup>3</sup> | 2 (0.3) | 34 (5.0) |
| **Father's Highest Level of Education [n (%)]** | **639<sup>4</sup>** | **677<sup>5</sup>** |
| Illiterate/No formal education | 210 (32.9) | 74 (10.9) |
| Primary (up to 5 Years) | 124 (19.4) | 84 (12.4) |
| Secondary/Matric/Class 9-10 | 175 (27.4) | 281 (41.5) |
| Intermediate (Class 11-12) | 65 (10.2) | 94 (13.9) |
| Graduate (14 years total education) | 58 (9.1) | 87 (12.9) |
| Professional degree<sup>3</sup> | 7 (1.1) | 57 (8.4) |

<sup>1</sup> Of 642 families with children under age 5, data missing for one mother; <sup>2</sup> Of 679 families with children under age 5, data missing for two mothers; <sup>3</sup> Beyond graduate, e.g. medicine, teaching, law, etc; <sup>4</sup> Of 642 families with children under age 5, data missing for one father and two fathers who were deceased; <sup>5</sup> Of 679 families with children under age 5, data missing for two fathers.

Another metric collected in Study 2 was the highest level of education achieved in the household given that the parents of children under age five were only a subset of household members. Data were available on 899 of 973 households. The median level of education was Class 12 (intermediate) with interquartile range Class 9-10 (secondary through matriculation), and Class 14 (bachelor’s or graduate). Of 899 households, 130 (14.5%) reported highest level as secondary through matriculation; 222 (24.7%) reported Class 12, 213 (23.7%) reported Class 14, 194 (21.6%) reported university master’s or above. Eight (including 5 included in other categories) reported professional training and other certificates such as Lady Health Worker, primary teaching certificate, and Montessori diploma. Few households had lower educational status: only 90 (10%) had Class 6-8 as the highest education, 43 (4.8%) had primary school, and 4 (0.4%) reported no education.

### 3.5 Occupations

Table 4 shows the occupations of parents of children under age five in both studies, and all occupations reported by household members who earned income. Supplementary Table 2 shows categories of occupation in Study 1 versus Study 2. Supplementary Table 3 shows the specific detailed occupations as categorized into High, Medium and Low perceived status.

**Table 4.** Occupations of Parents of Children under age 5 years in Study 1 and Study 2 and of all paid Household Members in Study 2.

| Occupational Category | Study 1<br>Parents of Children U5<br>n (%) n = 641<sup>1</sup> |  | Study 2<br>Parents of Children U5<br>n (%) n=677 |  | Study 2<br>All Household Members<br>Reporting Employment<br>n (%) n = 1697<sup>2</sup> |  |
| --- | --- | --- | --- | --- | --- | --- |
|  | Mother | Father<sup>3</sup> | Mother | Father<sup>3</sup> | Female<br>n = 246 (14.5) | Male<br>n = 1448<br>(85.5) |
| Unemployed | 6 (0.9) | 42 (6.6) | 1 (0.1) | 38(5.61) | 49 (19.9) | 161 (11.2) |
| Housewife | 577 (90.0) | 0 | 591 (87.3) | - | 591<sup>4</sup> | - |
| Laborer/Farmer | 1 (0.2) | 221 (34.5) | 0 | 61 (9.0) | 8 (3.3) | 119 (8.2) |
| Army Worker/Protective services | 0 | 100 (15.6) | 1( 0.1) | 123 (18.2) | 14 (5.7) | 238 (16.4) |
| Student | 16 (2.5) | 27(4.2) | 3 (0.4) | 3(0.4) | - | - |
| Office Worker | 13 (2.0) | 73 (11.4) | - | 11 (1.6) | 2 (0.8) | 24 (1.8) |
| Health Care | - | - | 8(1.2) | 11 (1.6) | 22 (8.9) | 34 (2.3) |
| Business | 0 | 100 (15.6) | 2(0.3) | 154 (22.8) | 15 (6.1) | 311 (21.5) |
| Skilled Worker | - | - | 9(1.3) | 171 (25.3) | 24 (9.8) | 366 (25.2) |
| Teacher | 27 (4.2) | 38 (5.9) | 54(8.0) | 34 (5.0) | 90 (36.6) | 61 (4.2) |
| Police (additional category in Study 1) | 0 | 10 (1.6) |  |  |  |  |
| Driver | 0 | 28(4.4) |  |  |  |  |
| Landlord | 1 (0.2) | 0 | - | 3 (0.4) | 0 | 1 (0.1) |
| Finance Related | - | - |  | 6 (0.9) | 0 | 12 (0.8) |
| Administration/Management/ Executive | - | - | 6 (0.9) | 41 (6.1) | 6 (2.4) | 60 (4.1) |
| Professional | - | - | 2 (0.3) | 20 (3.0) | 16 (6.5) | 61 (4.2) |

<sup>1</sup> Of 642 total families, occupational data were missing for one family; <sup>2</sup> n = 893 distinct households reported members with an occupation. Median number of individuals per household who work = 1 (IQR 1,2); <sup>3</sup> n = 2 fathers (0.3) reported as “Deceased” in occupation category in Study 1, one missing in Study 2; Data for 3 males missing in Study 2 <sup>4</sup> Housewives not counted amongst those employed for income; - Category not available for the related study.

In Study 1, there were only nine occupations reported. Most mothers of children under five years (n=577/641, 90%) were housewives. A few were teachers (n = 27, 4.2%), office workers (n= 13, 2%) or students (n = 16, 2.5%). In Study 2, the majority, (n=591/677, 87.3%) were housewives. However, there was a greater variety of professions besides teaching (n=54, 8.0%), including health care workers (n=8, 1.2%), administrative, management and executive roles (n=6, 0.9%), professionals including a physical scientist and economist (n=2, 0.3%), and those doing tailoring, hairdressers/beauticians, housekeeping and related jobs (9, 1.3%).

In Study 1, the most common occupations for fathers of children under five years were laborer/farmer (n=221/6412, 34.5%), followed by business and army (n=100 each, 15.6%), office workers (n=73, 11.4%), teachers (n=38, 5.9%), and students (n=27, 4.2%); 42 (6.6%) reported being unemployed and 2 were deceased. In Study 2, of 676 fathers, a much smaller number were laborers/farmers (61, 9%) than for Study 1. The proportion in the army and those who were teachers remained about the same. A higher percentage were in business and in the skilled worker category, both categories having a wide variety of possible jobs. Forty-one (6.1%) reported being in the administrative, management or executive category, and 20 (3%) were in the professional group.

In Study 2, 1696 individuals from 893 households reported occupation data. Households had a median of one individual employed (IQR 1,2, range 0-7). Fifty-nine different occupations were reported. In addition to 121 retirees (7.1%), 89 were unemployed (5.3%). Most of those employed were men (1448, 85.5%). The most frequent occupations for men were working properties (e.g., shopkeeper and businessman) (n=300, 17.7%), skilled workers (366, 25.2%) army and protective service workers (n=238, 16.42.3%), laborer/farmer (119, 8.2%), professionals 61 (4.2%), teachers (61, 4.2%), administrative/management and executives (60, 4.1%), and unskilled labor (110, 6.5%). Sixty individuals (3.5%) reported a second occupation. For the 246 females who reported paid employment, the most common categories were teachers (90, 36.6%), skilled workers (24, 9.8%), health care (22, 8.9%), professionals (6.5%), business (15, 6.1%), and army or protective services (5.7%). The median monthly income of households in 2012 PKR was 22000 (IQR 12000, 40000). The annual agricultural income was 22000 (IQR 10000, 50000).

### 3.5 Socioeconomic Status

The exploratory factor analysis of Study 1 revealed two factors, one composed of variables describing the construction of houses and the second describing access to water and sanitation (**Table 3**). The two factors explained approximately half the variance in the sample (28 and 22%, respectively). This two-factor structure was a reasonable fit to the Study 2 SES data (Tucker-Lewis index 0.98, RMSEA 0.01) but explained a little less variance in the data (23.4% and 18%, respectively).

**Table 5:**
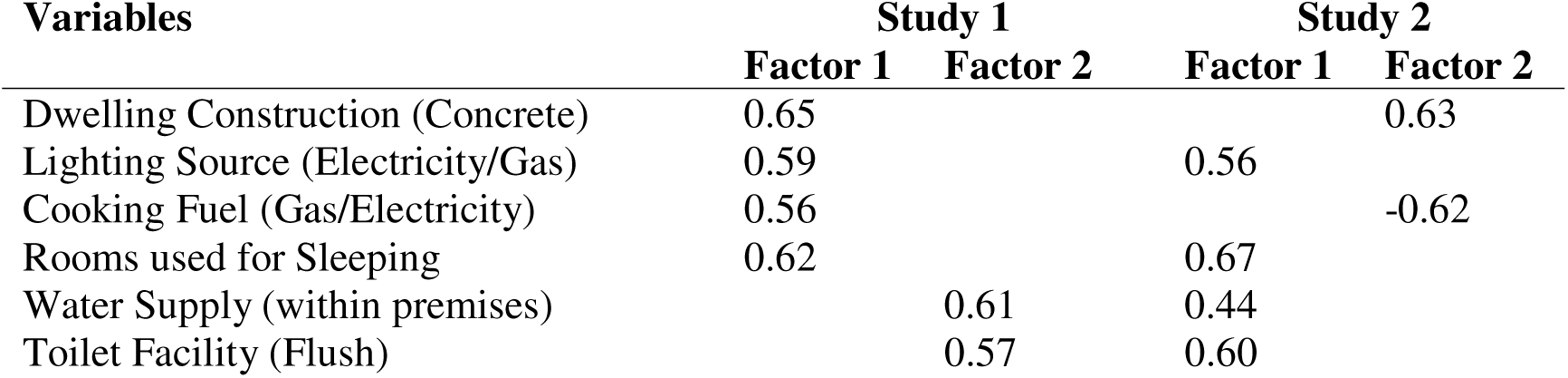
Factor loadings of Socioeconomic Status from Study 1 and Study 2.

| Variables | Study 1 |  | Study 2 |  |
| --- | --- | --- | --- | --- |
|  | Factor 1 | Factor 2 | Factor 1 | Factor 2 |
| Dwelling Construction (Concrete) | 0.65 |  |  | 0.63 |
| Lighting Source (Electricity/Gas) | 0.59 |  | 0.56 |  |
| Cooking Fuel (Gas/Electricity) | 0.56 |  |  | -0.62 |
| Rooms used for Sleeping | 0.62 |  | 0.67 |  |
| Water Supply (within premises) |  | 0.61 | 0.44 |  |
| Toilet Facility (Flush) |  | 0.57 | 0.60 |  |

The SES scores for new households that moved into the village by Study 2 were on average significantly lower than the score of original households in Study 1 (Study 2, mean –0.47, SD 0.144; Study 1 mean 0.16, SD 0.1; T-test, p = 0.001) (**Figure 3**). The difference between original and newer households was narrowed with the additional variables in Study 2 (**Figure 3**).

**Figure 3:**
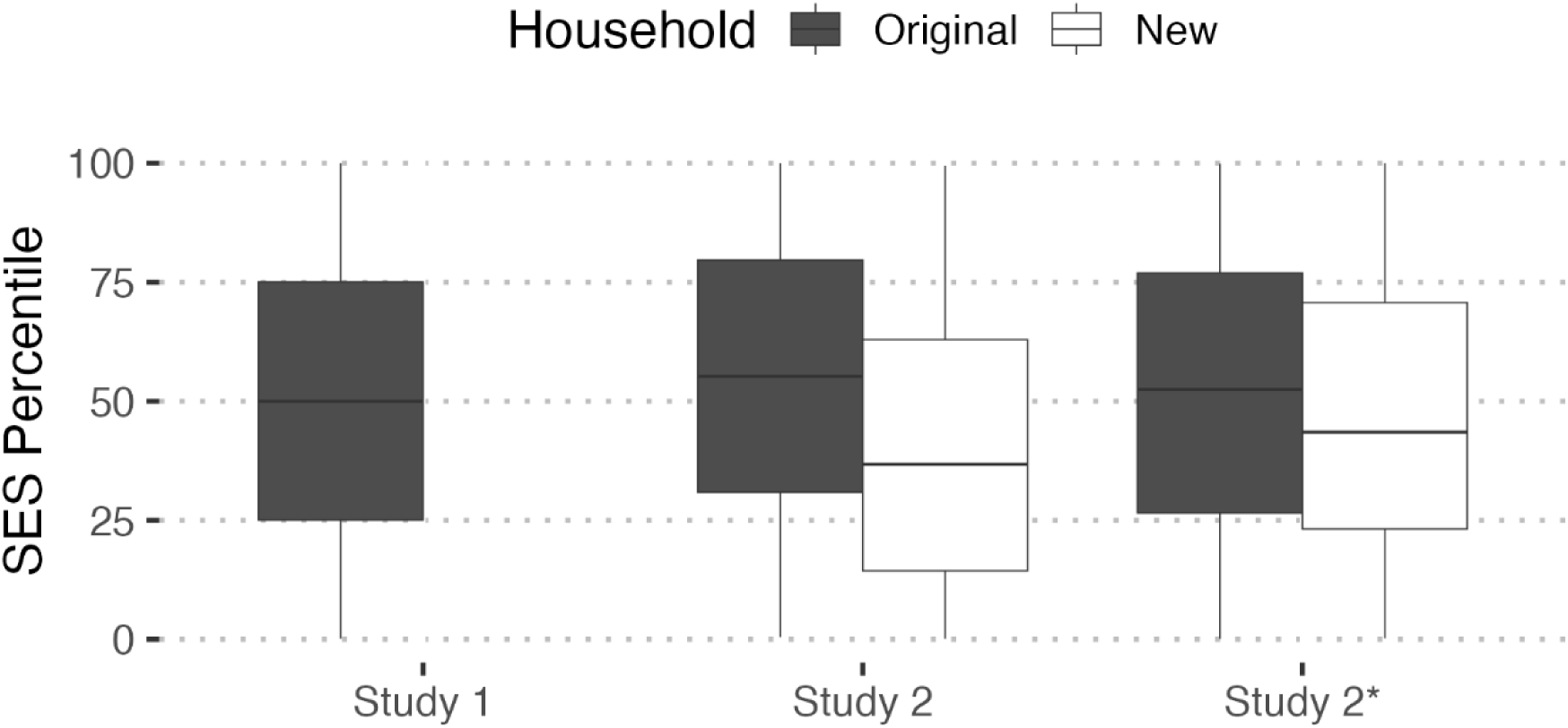
Distribution of socioeconomic status percentiles of original households and new households in the population at Study 1 and Study 2 using common variables and then using the wider set of variables specific to Study 2(*).

Households tended to move equally between quintiles from Study 1 to Study 2. Roughly the same proportion increased (mean 22%, range 16.6 to 33.3%), stayed the same (mean 21.7%, range 18 to 24%) or decreased (mean 16.6%, range 3.8 to 27%) their quintile (**Figure 4**). The smallest change was from middling SES in Study 1 to the lowest quintile in Study 2. Households that moved from lowest to highest quintiles tended to have lower household density from Study 1 to Study 2 (5.7 to 3 people per room), have fourfold higher mean monthly savings in Study 2 (from 1714 to 7143 Pakistani Rupees), and a 2.7-fold higher mean land holdings (160 to 434 marlah, where 1 marlah is approximately 25.3 m²). Households moving from the highest to lowest quintiles typically had similar differences in the opposite direction.

**Figure 4:**
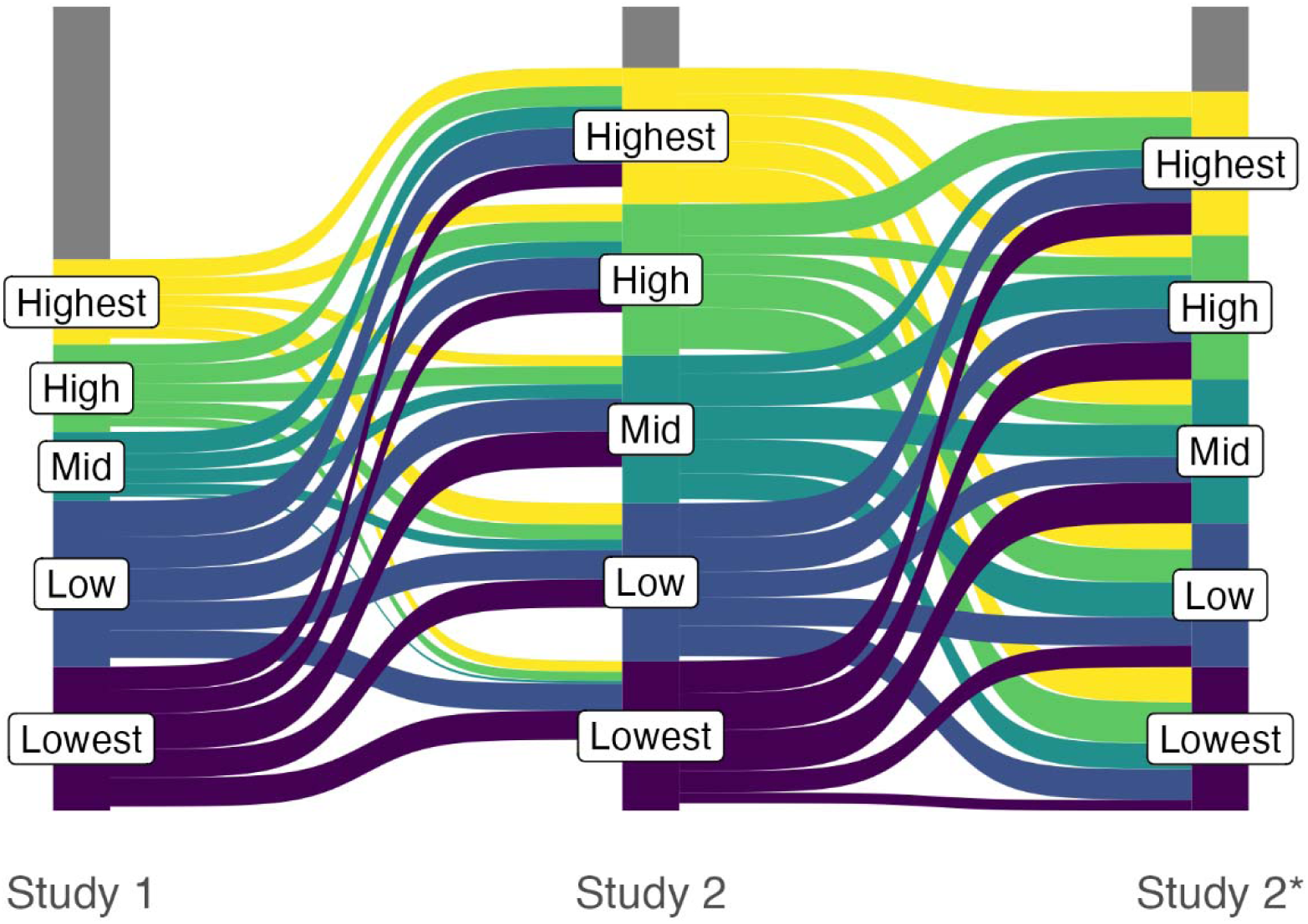
Change in the quintile of socioeconomic status between Study 1 and Study 2 using common variables and then using the wider set of variables specific to Study 2(*). Grey bars indicate the different number of households at each timepoint or that had missing data.

Constructing a bespoke SES variable for Study 2 using all available information resulted in seven factors that were retained from 55 original variables (See Supplementary Table 1).

The expanded range of variables included information on assets that indicate short-term (e.g., the number of household members earning an income) and long-term (e.g., ownership of agricultural land and savings) wealth. Of the variables available, the type of construction, location of stove (by season), presence of a metal chimney at home, the type of ventilation in the kitchen, and presence of smoke in the household did not load onto any factor.

**TABLE 6.**
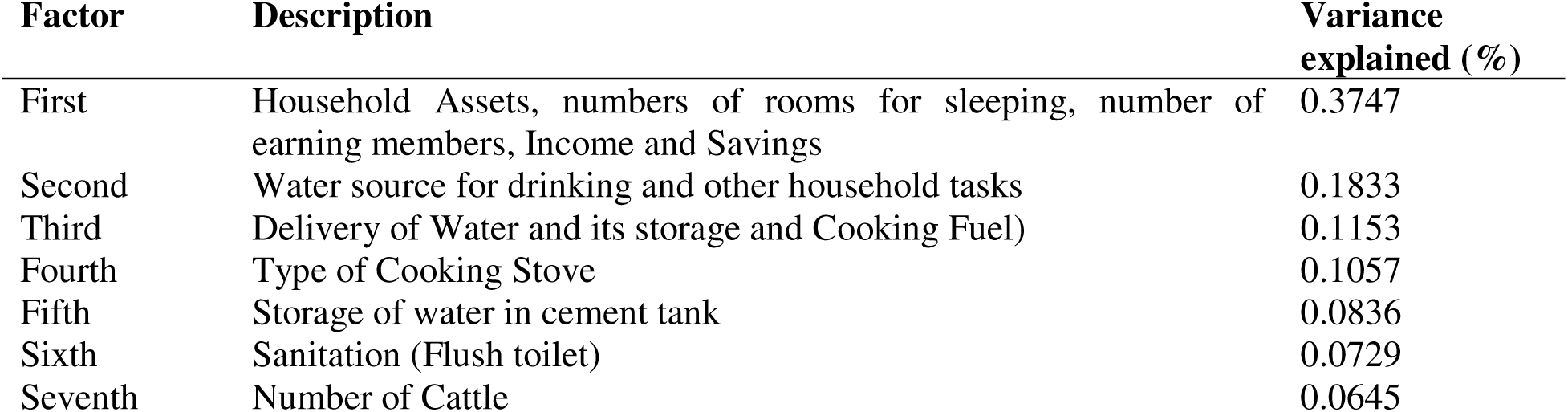
Description of factor describing Socioeconomic Status using data from Study 2.

| Factor | Description | Variance explained (%) |
| --- | --- | --- |
| First | Household Assets, numbers of rooms for sleeping, number of earning members, Income and Savings | 0.3747 |
| Second | Water source for drinking and other household tasks | 0.1833 |
| Third | Delivery of Water and its storage and Cooking Fuel) | 0.1153 |
| Fourth | Type of Cooking Stove | 0.1057 |
| Fifth | Storage of water in cement tank | 0.0836 |
| Sixth | Sanitation (Flush toilet) | 0.0729 |
| Seventh | Number of Cattle | 0.0645 |

There was no statistically significant pattern between household salaries and household SES rank (ANOVA, p = 0.2) though there was a significant, but small effect of agricultural income (mean 0.02 increase in SES rank per 1000 PKR, p = 0.046). Similarly, of those households with livestock, SES ranks tended to be higher with the more animals owned, in particular for cattle and poultry (**Figure 5**, SES percentile increased 0.8 per additional animal, p <0.001) but not the amount of agricultural land (ANOVA, p = 0.37).

**Figure 5:**
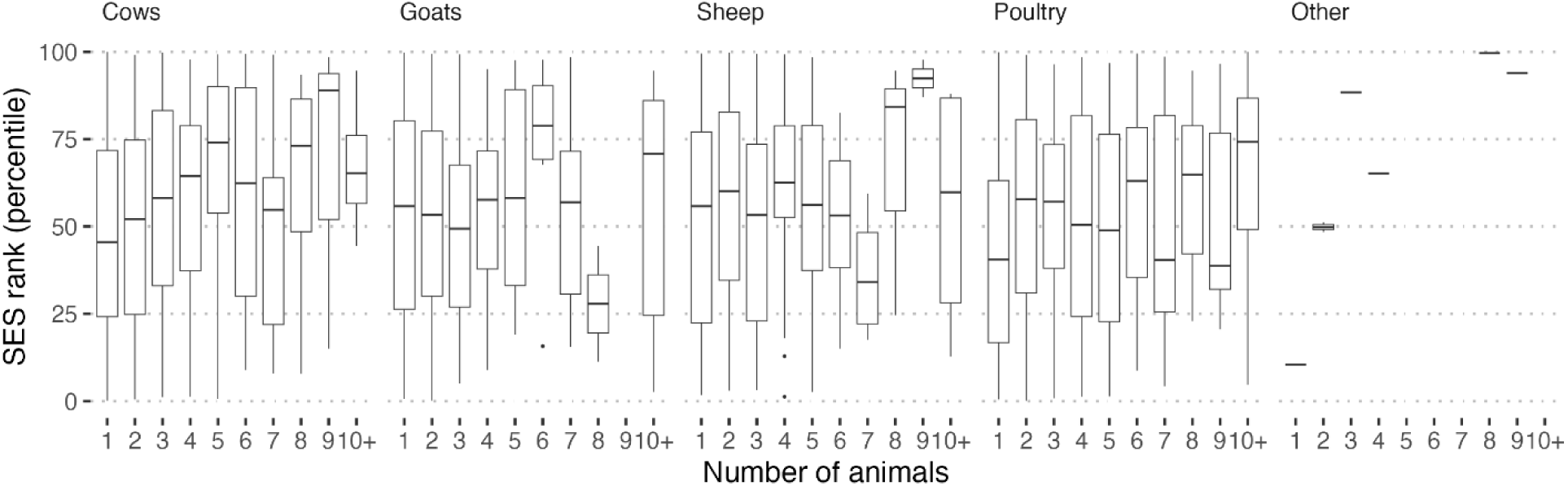
Socioeconomic rank given the number of animals owned in Study 2.

Lower status jobs tended to be associated with lower household SES (mean SES percentile 47.5) compared to medium or higher status jobs (51.1 or 52.1 respectively) (ANOVA, p = 0.02). Different occupations, but not their classified status (high/medium/low), were strongly associated with household SES ranks (**Figure 6**). Salaries were similarly associated with job status and like the SES association, middle and higher status occupations had similar mean salaries (median 40000 PKR, IQR 25000 to 68000 and 38000 PKR, 20000 to 71000, respectively) that were both higher than low status occupations (20500 PKR, IQR 12000 to 40000).

**Figure 6:**
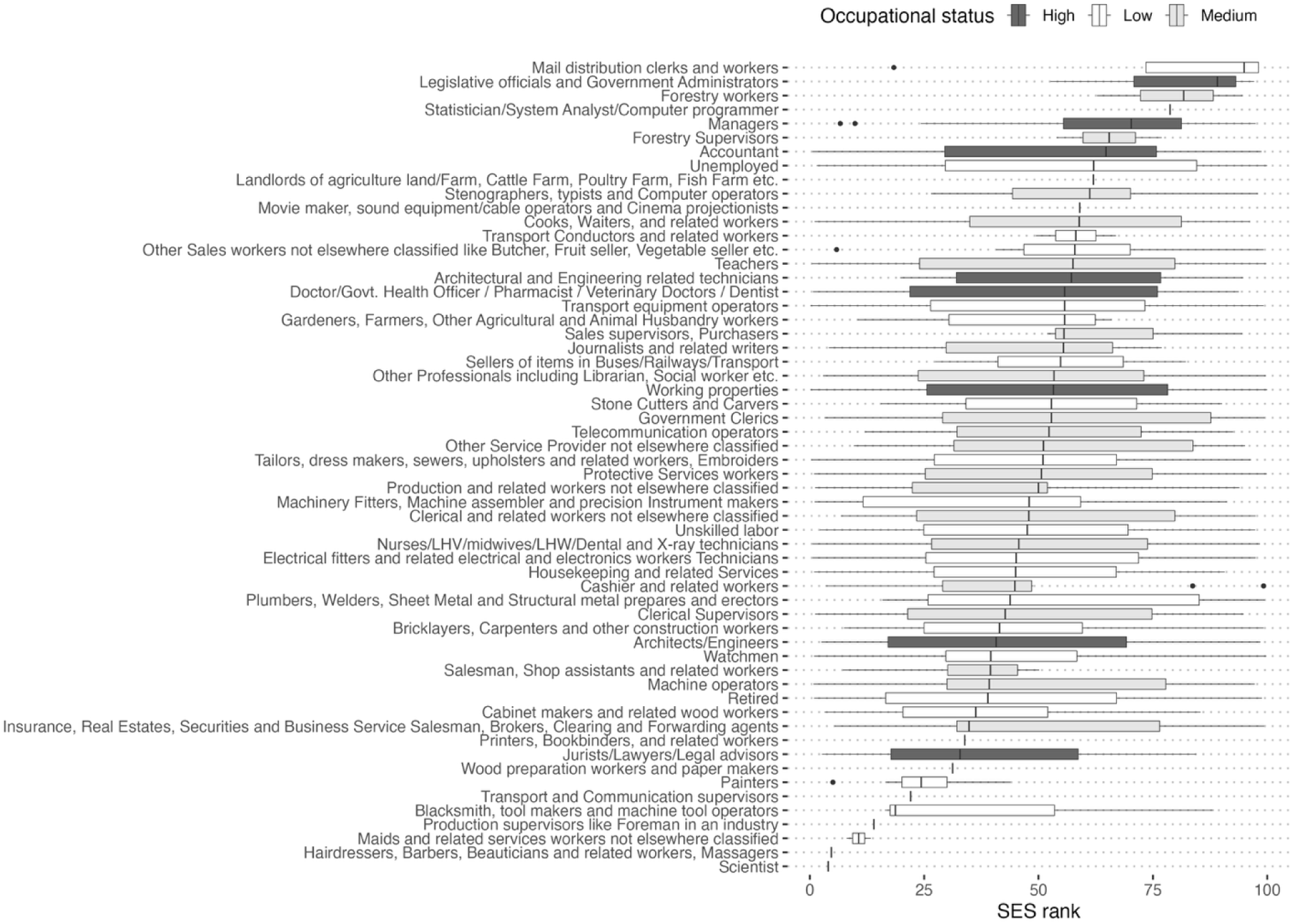
Household socioeconomic rank by different occupations. Occupations were manually classified by perceived status (high, dark grey; medium, light grey; low, white).

## 4 Discussion

SES is an important driver of health outcomes and human capital – the realization of human productivity. We report on the transitions within a remote and rural village over two decades that has out-paced national trends and signifies the success of targeted investments, particularly in female education and family planning, water and sanitation. This population is in a remote area of Pakistan, in a country that ranks low on the Human Development Index. We report dramatic changes including the transformation from a typically low-income demographic distribution to a middle-income distribution, increased employment and educational opportunities, especially for women. Self-reported illnesses were relatively rare and unrelated to SES.

Recognition of the importance of poverty in this strategically important area of Pakistan led to targeted investments since the 1980s that have resulted in socioeconomic developments outpacing the national trends and which demonstrate the success of these initiatives. Investments in this population in the early 1990s, in particular electrification through hydroelectric power projects and the construction of girls’ schools dramatically changed the outlook of this population (World Bank 2010, Benz, 2013). By Study 2, all households had access to electricity even if the supply was still seasonally inconsistent. Construction of homes improved with half having cement/brick structures versus the earlier period, where a third had mud/stone only. Households had also diversified their back-up fuel sources with, for example, rechargeable electric lights and gas, with the elimination of kerosene. Together these changes are important because they have been shown to relate to lower levels of respiratory disease (Aftab A, et al., 2022) and greater stability in natural disasters, e.g., landslides (Smith, R.B., 2023), and better comfort in adverse weather.

Similarly, investment in water filtration plants was matched with a change in access to clean water and household sanitation as households moved away from traditional methods of extracting water from glacial streams and many could pipe water directly into the home; instead of just letting water settle as in Study 1, most families who treated water in Study 2 boiled it. Traditional outhouse toilets were largely replaced by flush ones. Consequently, child mortality from diarrheal diseases dropped (Hansen et al., 2020).

The average household size by Study 2 was comparable to that of national averages, with 6.18 persons for rural areas (Pakistan Bureau of Statistics, 2026). Methods of cooking in the household lagged behind other developments, and the majority of households still relied on wood for cooking in Study 2, although many had moved kitchens into the home and installed a chimney. Wood continued to be the main source of heating in this area which has cold temperatures in winter. It was common to see a haze of smoke above the village during winter. The finding that more than 20% of households had smokers who mainly smoked inside the house was also a finding of concern. Indoor pollution has remained a challenge to respiratory health in this and similar populations (Liaqat et al, 2025, Colbeck et al, 2010).

The demographic pyramid in the early 1990s was typical of a high fertility, high mortality, low-income population (“stage 1”) (Pakistan Demographic and Health Survey 1990-1991, 1992). Within just 20 years that demographic pattern changed to a pattern indicative of a middle-income population with a halving of the proportion of children under five and consequently a lower dependency ratio between adults of working age and the very young or old (“stage 3”). This contrasts with the national demographic pyramid that remains in Stage 1 (Pakistan Demographic and Health Survey 2012-13, 2013, Pezzulo et al, 2017, United Nations, 2024). The Oshikhandass population has achieved a distribution closer to that in developed countries sooner than Pakistan as a whole. It is likely that the introduction of family planning services in the late 1990s contributed significantly to this change (Aga Khan Health Service, 1998) by enabling women to have the family size they desired. The demographic transition away from high fertility to compensate for high mortality (particularly infant mortality) was matched by an economic transition with changing employment opportunities moving from a predominantly agricultural setting to an increasingly urban population structure. Many people were able to find employment in Gilgit. Of note, however, the village continued to appear rural and wealthier households tended to have income from land. We observed occupations changing from predominantly agricultural occupations for men and women, who were mainly housewives in Study 1, to a wide range of occupations in Study 2 including diversification into services, administration and retail.

We found very high levels of education amongst parents of children under age 5 years, especially in mothers, in Study 2 compared with Study 1. This is substantially higher than the Pakistani average literacy rate in 2024-2025 of 73% for males and 54% for females (HIES). It is evident that within one generation, women with children under age 5 were becoming highly educated, reflecting the substantial investments made in girls’ education during the same period. The even more remarkable finding was the high level of education in households in Study 2: 629/899 (70%) of households had someone with Class 12 (intermediate) or higher education, with 213 (24%) having a college degree and 194 (22%) a university master’s degree or higher. In comparison, the national rates of those completing upper secondary level (Class 12) in Pakistan in 2021-2022 was 23% (Pakistan Economic Survey 2023-2024). These results concur with national-level modelling that highlight the synergistic effects of investing in skills as well as the physical environment to promote development of human capital (Tam, 2024).

At face value, the relative SES score of households that had been in Study 1 remained relatively similar in Study 2 using characteristics of the house structure. Despite having access to improved construction materials and being able to access improved water and sanitation, the newer households in the village still tended to have lower SES ranks. This is likely due to newer families coming from poorer neighboring villages in order to access opportunities in Oshikhandass, which also benefits from closer proximity and better access to the administrative capital, Gilgit. The increased land and incomes of households showing the greatest improvements in SES acted as draw factors to poorer surrounding villages (Ullah et Al, 2024). The addition of variables describing the people (i.e., occupation, income) and not just household structures narrowed this difference, highlighting how different aspects of SES can lead to different conclusions about socioeconomic position within a population (Zaneva et al, 2024). There could also be factors which pulled families from Oshikhandass to other areas. While we did not collect data on those who had migrated out after 1996, a follow up study done of adolescents and young adults (Rasmussen et al., 2021) indicated that of those who moved, more than half had done so for continued educational opportunities in larger cities.

The change in SES between the two study periods showed increases and decreases in approximately equal frequency. It might be expected that households reassort their relative socioeconomic position (National Academies of Sciences, Engineering and Medicine, 2022), but it is surprising to see so many decreases in SES given the consistent improvement in, for example, housing stock. To illustrate, households that increased from the lowest to highest quintiles tended to increase the number of rooms used for sleeping, noting that these would not necessarily be dedicated bedrooms. Access to electrical backup generators was also a feature of households that change quintile, either gaining access and improving or losing access to these expensive resources and decreasing SES quintiles. The addition of more variables in Study 2 gives a richer interpretation of SES, but the difference in SES ranks between the confirmatory and exploratory analyses of the Study 2 data still showed frequent decreases in relative position including from the highest to lowest quintiles and vice versa. For example, changes in quintile were also associated with income and land that were not recorded in Study 1, but these captured important trends in how SES might manifest. Many different operationalization of SES exist (Zaneva, 2024) Even in our analysis of similar variables, relative SES substantially changed over time. One solution is to use subjective assessments of SES rather than attempting to gather objective indicators (Prag, 2020), which has the advantage of being contextualized to a population but the disadvantage of comparing between populations (or over time).

We only found a weak association between occupational status and socioeconomic position, the former based on classifying occupations perceived to be high, medium and low. This was somewhat surprising because incomes were strongly associated with occupational status. Household incomes were, however, the aggregate of all household members. Not all jobs fit the expected pattern; notable exceptions were for postal workers who tended be from high-SES households despite low status jobs, and conversely lawyers who had the reverse pattern. On further investigation, postal workers lived in households with other sources of income besides employment, such as agricultural income, and had other assets. Of the 2 lawyers, one did not report individual income, so appeared to have low status.

SES is reportedly a strong predictor of health outcomes, but the studies of Oshikhandass focused on childhood illness showed that was not strongly related to aggregate indices of SES (Hansen et al., 2020), but did relate to specific aspects, particularly parental education and language (Rasmussen et al., 2021). Fewer indicators of adult morbidity were recorded and there were proportionately few deaths.

Consequently, there were no statistically significant associations between self-reported adult chronic disease and SES. This study was not designed to assess linkages between adult illness and SES. If that had been intended, it would have been important to obtain additional information from medical records maintained in the household, or local clinics or at nearby hospitals. It would also have been important to have objective indicators of illness, such as measurement of weight, abdominal girth, blood pressure, blood sugar, lipids and liver function tests, and respiratory status to assess for chronic diseases such as overweight/obesity, hypertension, diabetes mellitus, hyperlipidemia, metabolic dysfunction-associated steatotic liver disease, and chronic lung disease. Mulyanto et al (2019) note that the use of self-report may lead to underestimation of socioeconomic inequalities in disease prevalence and recommend use of objective assessment. This underlines the limitations of self-report and emphasizes the complexity of studies for population-based chronic disease, which was beyond the scope of this study.

Based on what we learned in this study, community-based and potentially other measures (e.g., health facility, workplace) to decrease smoking and indoor air pollution would be important. Based on death data noting that hypertension, cardiac disease, cancer, and diabetes were common causes of death, and on data from those alive which indicate that the same conditions were common, community-based efforts to screen for these would be useful. Additionally, poor mental health was a cause of morbidity and is under-recognized in similar populations (Mumford et al., 1996).

## LIMITATIONS

Neither Study 1 nor Study 2 were designed to collect and characterize SES other than to control for factors relating to health outcomes. As such the variables collected in Study 2 were not all available in Study 1 because appreciation of the factors that might influence the two health outcomes (childhood diarrhea and pneumonia) changed as new research became available. We attempted to control for variables that were consistent and to provide a narrative description of broader changes in the population. A further significant limitation to our study was that all health conditions were by self-report and not through measurement or documentation by medical records. This undoubtedly results in under-reporting of disease, given that many, like hypertension, are not accompanied by symptoms. Rasmussen et. al. (2021) reported that in adolescents and young adults from the same population who had blood pressure measured, 12.7% had Stage 1 or 2 hypertension and 2.9% had elevated blood pressure. We could not assess relationship of health status to socioeconomic status given limitations in measurements of health status.

## CONCLUSIONS

We describe a community which is primarily a success story about targeted investments and how over a single generation a population transformed from a predominantly low-income agricultural community to a middle-income more urban setting. The proliferation of opportunities has been particularly pronounced for women. The development of this population has been faster than the rest of the nation. Despite clear secular improvements in the whole population, individual SES ranks changed substantially using the metrics we used. Relative SES decreased as much as improved and the increased availability of data to capture different dimensions of SES demonstrate the need for caution in focusing on a limited number of metrics.

## Data Availability

All data produced in the present study are available upon reasonable request to the authors. Deidentified data have been uploaded to Zenodo with DOI 10.5281/zenodo.18299618

https://zenodo.org/records/18299618

## Acknowledgements

Lena Schelzig, for assistance with literature searches, analysis and references; Chelsea L Hansen, for assistance with data curation and analysis; Faran Sikandar, for assistance in conceptualizing the study; Dr. Khalil Ahmed, for assistance in implementation; to all families and respondents in Oshikhandass who participated in the study.

