## Supplemental Table 2 for "Rapid changes in population socioeconomic status indicators are unevenly distributed in a rural Pakistani village"

Supplementary Table 2 Occupation Categories Study 1 vs Study 2

Key: Lavendar is Study 1 Categories; Green Denotes Additional Categories in Study 2

Classified/Perceived SES status is in parentheses, 1-3: 1 being highest SES, 2 being medium and 3 being lowest

| **Categories** | **SES** |
| --- | --- |
| **Unemployed** | - Retired - Unemployed |
| **Student** |  |
| **Housewife** |  |
| **Office Worker** | - Stenographers, typists and Computer operators (2) - Mail distribution clerks and workers (3) - Telecommunication operators (2) - Clerical and related workers not elsewhere classified (2) |
| **Teacher** |  |
| **Professional** | - Other Professionals including Librarian, Social worker etc. (2) - Journalists and related writers (2) - Scientist (Non-Biological) (1) - Architects/Engineers (1) - Architectural and Engineering related technicians (1) - Statistician/System Analyst/Computer programmer (1) - Accountant (1) - Jurists/Lawyers/Legal advisors (1) |
| **Finance Related** | - Cashier and related workers (2) |
| **Administration Management/**  **Executive** | - Legislative officials and Government Administrators (National and Provincial Assembly members, (GoP’s Grade 17 and above officials) (3) - Managers (Including Directors, General Managers, Administrative Managers) (3) - Clerical Supervisors (2) - Transport and Communication supervisors (2) - Government Clerks (Government of Pakistan’s Grade less than 17) - Production supervisors like Foreman in an industry (2) |
| **Businessman** | - Working properties (shopkeeper, Businessman) (1) - Sales supervisors, Purchasers (2) - Insurance, Real Estates, Securities and Business Service Salesman, Brokers, Clearing and Forwarding agents (2) - Salesman, Shop assistants and related workers (2) - Sellers of items in Buses/Railways/Transport (3) - Other Sales workers not elsewhere classified like Butcher, Fruit seller, Vegetable seller etc. (3) |
| **Laborer/Farmer** | - Transport Conductors and related workers (3) - Gardeners, Farmers, Other Agricultural and Animal Husbandry workers (3) - Forestry Supervisors (2) - Forestry Workers (2) - Maids and related services workers not elsewhere classified (3) - Unskilled labor |
| **Landlord** | - Landlords of agriculture land/Farm, Cattle Farm, Poultry Farm, Fish Farm etc. (1) |
| **Army Worker** | - Watchmen (3) - Protective Services workers (Fire fighters, Police, Army personnel’s etc.) (2) |
| **Health Care** | - Doctor/Govt. Health Officer/Pharmacist/Veterinary Doctors/Dentist (1) - Nurses/LHV/midwives/LHW/Dental and X-ray technicians (2) |
| **Skilled Worker** | - Movie maker, sound equipment/cable operators and Cinema projectionists (2) - Photographers/Calligraphers/Graphic designers/Textile designers/Fashion designers/Commercial artists (2) - Wood preparation workers and paper makers (3) - Tailors, dress makers, sewers, upholsters and related workers, Embroiderers (3) - Painters - Production and related workers not elsewhere classified - Cabinet makers and related wood workers (3) - Bricklayers, Carpenters and other construction workers - Stone Cutters and Carvers (3) - Machine operators - Transport equipment operators (Drivers, Sellers etc.) - Blacksmith, tool makers and machine tool operators (3) - Hairdressers, Barbers, Beauticians and related workers, Massagers (3) - Housekeeping and related Services (3) - Other Service Provider not elsewhere classified (2) - Cooks, Waiters, and related workers (2) - Machinery Fitters, Machine assembler and precision Instrument makers (except electrical) including Auto Mechanics (3) - Electrical fitters and related electrical and electronics workers Technicians (3) - Plumbers, Welders, Sheet Metal and Structural metal prepares and erectors - Printers, Bookbinders, and related workers (press workers) |
| **Deceased** |  |
