## Supplemental Table 4 for "Rapid changes in population socioeconomic status indicators are unevenly distributed in a rural Pakistani village"

Supplementary Table 4: Factor loadings of Socio-Economic Status from Study 2 Variables only

| **Variables** | **Factor 1** | **Factor 2** | **Factor 3** | **Factor 4** | **Factor 5** | **Factor 6** | **Factor 7** |
| --- | --- | --- | --- | --- | --- | --- | --- |
| 1. Dwelling Construction (Concrete Roof) |  |  |  |  |  |  |  |
| 2. Dwelling Construction (Wooden Roof) |  |  |  |  |  |  |  |
| 3. Exterior Wall (Brick, Cement or Concrete) |  |  |  |  |  |  |  |
| 4. Lighting Source (Electricity/Gas) |  |  |  |  |  |  |  |
| 5. Rooms used for Sleeping | 0.5580 |  |  |  |  |  |  |
| 6. Heating Source (Electricity/Gas) |  |  |  |  |  |  |  |
| 7. Type of Cooking Stove (BACIP/Electric/Gas) |  |  |  | 0.5585 |  |  |  |
| 8. Cooking Fuel (Electricity/Gas) |  |  | 0.5930 |  |  |  |  |
| 9. Had Metal Chimney at home |  |  |  |  |  |  |  |
| 10. Location of Stove during Summer |  |  |  |  |  |  |  |
| 11. Location of Stove during Winter |  |  |  |  |  |  |  |
| 12. Type of Kitchen Ventilation (BACIP roof hatch/Exhaust) |  |  |  |  |  |  |  |
| 13. Had smoke in the house |  |  |  |  |  |  |  |
| 14. Toilet Facility (Twin Pit Latrine) |  |  |  |  |  |  |  |
| 15. Toilet Facility (Flush to Septic Tank) |  |  |  |  |  | -0.6184 |  |
| 16. Toilet Facility (Flush to Pit latrine) |  |  |  |  |  | 0.5945 |  |
| 17. Access to Water Filtration Plant |  | -0.6533 |  |  |  |  |  |
| 18. Main Source of Water for Household Task (Dar Filter Plant) |  | -0.7105 |  |  |  |  |  |
| 19. Water Supply (piped within premises) | 0.5491 |  |  |  |  |  |  |
| 20. Storage of Water in Plastic Tank for household Task | 0.3699 |  |  |  |  |  |  |
| 21. Storage of Water in Cement Tank for household Task |  |  | -0.5873 |  |  |  |  |
| 22. Main source of drinking water for the household (Dar Filter Plant/Bottled water) |  | -0.6281 |  |  |  |  |  |
| 23. Delivery of water at home for drinking (Piped to dwelling/plot) | 0.5109 |  |  |  |  |  |  |
| 24. Source of drinking water for the household when the main source is not available (Dar Filter Plant/Bottled water) |  |  |  |  |  |  |  |
| 25. Delivery of water at home for drinking when the main source is not available (Piped to dwelling/plot or buying) |  |  |  |  |  |  |  |
| 26. If Piped water, is it continuous or interrupted (Continuous) |  | 0.4000 |  |  |  |  |  |
| 27. Water Filter/Filter Machine at home as usual treatment (Water Filter) |  |  |  |  |  |  |  |
| 28. Boil Water as usual treatment (Boil Water) |  |  |  |  |  |  |  |
| 29. Storage of Water in Plastic Tank for drinking | 0.3496 |  |  |  |  |  |  |
| 30. Storage of Water in Cement Tank for drinking |  |  | -0.5930 |  |  |  |  |
| 31. Had Iron | 0.3732 |  |  |  |  |  |  |
| 32. Had Bed | 0.5156 |  |  |  |  |  |  |
| 33. Had Chair | 0.5320 |  |  |  |  |  |  |
| 34. Had Sofa | 0.3970 |  |  |  |  |  |  |
| 35. Had Cupboard | 0.5614 |  |  |  |  |  |  |
| 36. Had Table | 0.5661 |  |  |  |  |  |  |
| 37. Had Electric Fan |  |  |  |  |  |  |  |
| 38. Had Radio |  |  |  |  |  |  |  |
| 39. Had Computer | 0.4867 |  |  |  |  |  |  |
| 40. Had TV | 0.4323 |  |  |  |  |  |  |
| 41. Had Mobile Phone |  |  |  |  |  |  |  |
| 42. Had Refrigerator | 0.4481 |  |  |  |  |  |  |
| 43. Had Freezer |  |  |  |  |  |  |  |
| 44. Had Watch/Clock | 0.4570 |  |  |  |  |  |  |
| 45. Had Bank Account | 0.4934 |  |  |  |  |  |  |
| 46. Had Motor Vehicle | 0.3699 |  |  |  |  |  |  |
| 47. Had Cow (actual numbers) | 0.5066 |  |  |  |  |  |  |
| 48. Had Goat (actual numbers) |  |  |  |  |  |  |  |
| 49. Had Sheep (actual numbers) |  |  |  |  |  |  |  |
| 50. Had Chicken (actual numbers) |  |  |  |  |  |  |  |
| 51. Had Duck (actual numbers) |  |  |  |  |  |  |  |
| 52. Number of members earner in the household | 0.4241 |  |  |  |  |  |  |
| 53. Total Monthly Income from all sources | 0.4879 |  |  |  |  |  |  |
| 54. Annual Income from Agriculture |  |  |  |  |  |  |  |
| 55. Total monthly savings | 0.3387 |  |  |  |  |  |  |
| 56. Total Land overall (in Marlah) |  |  |  |  |  |  |  |

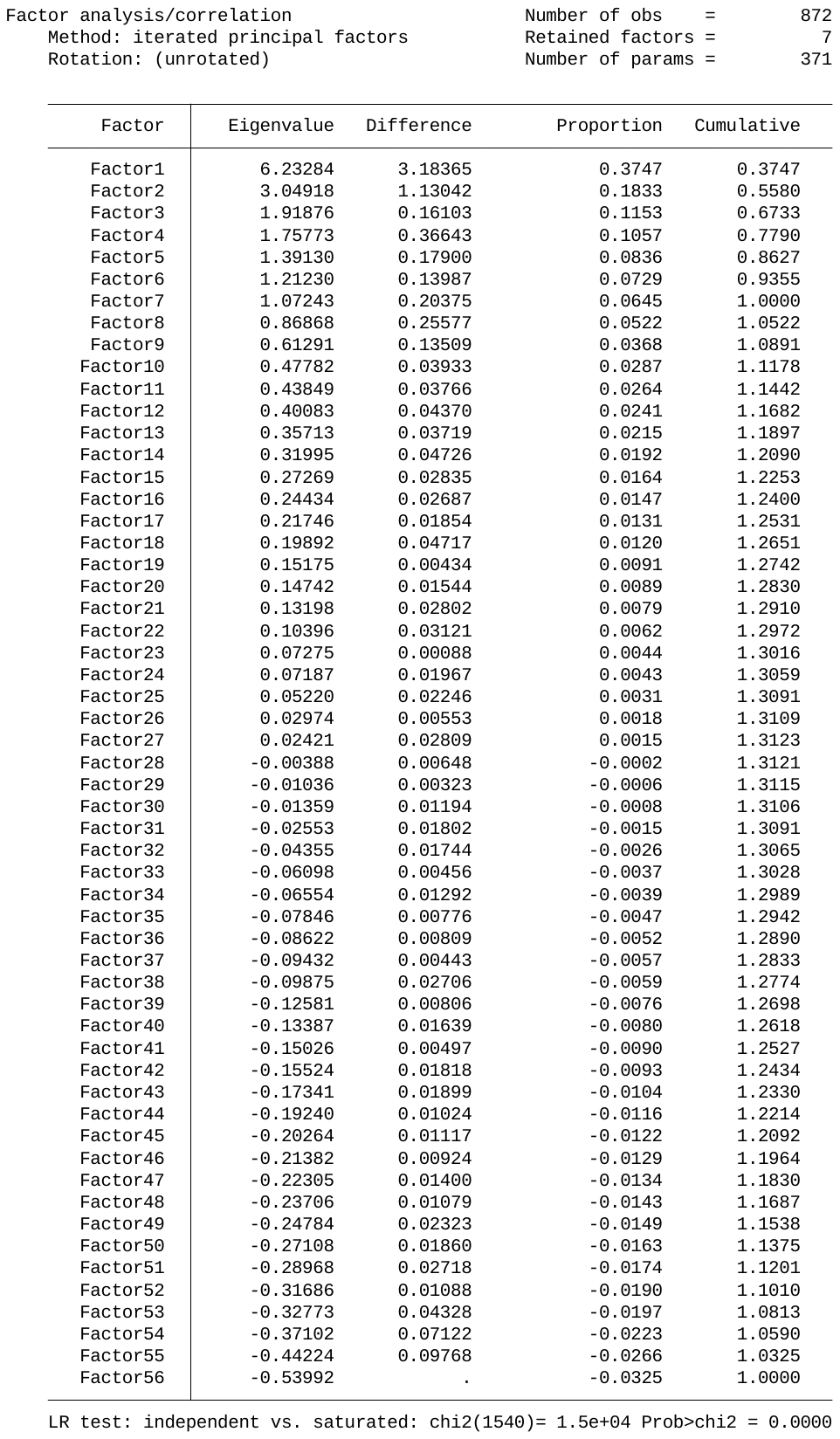

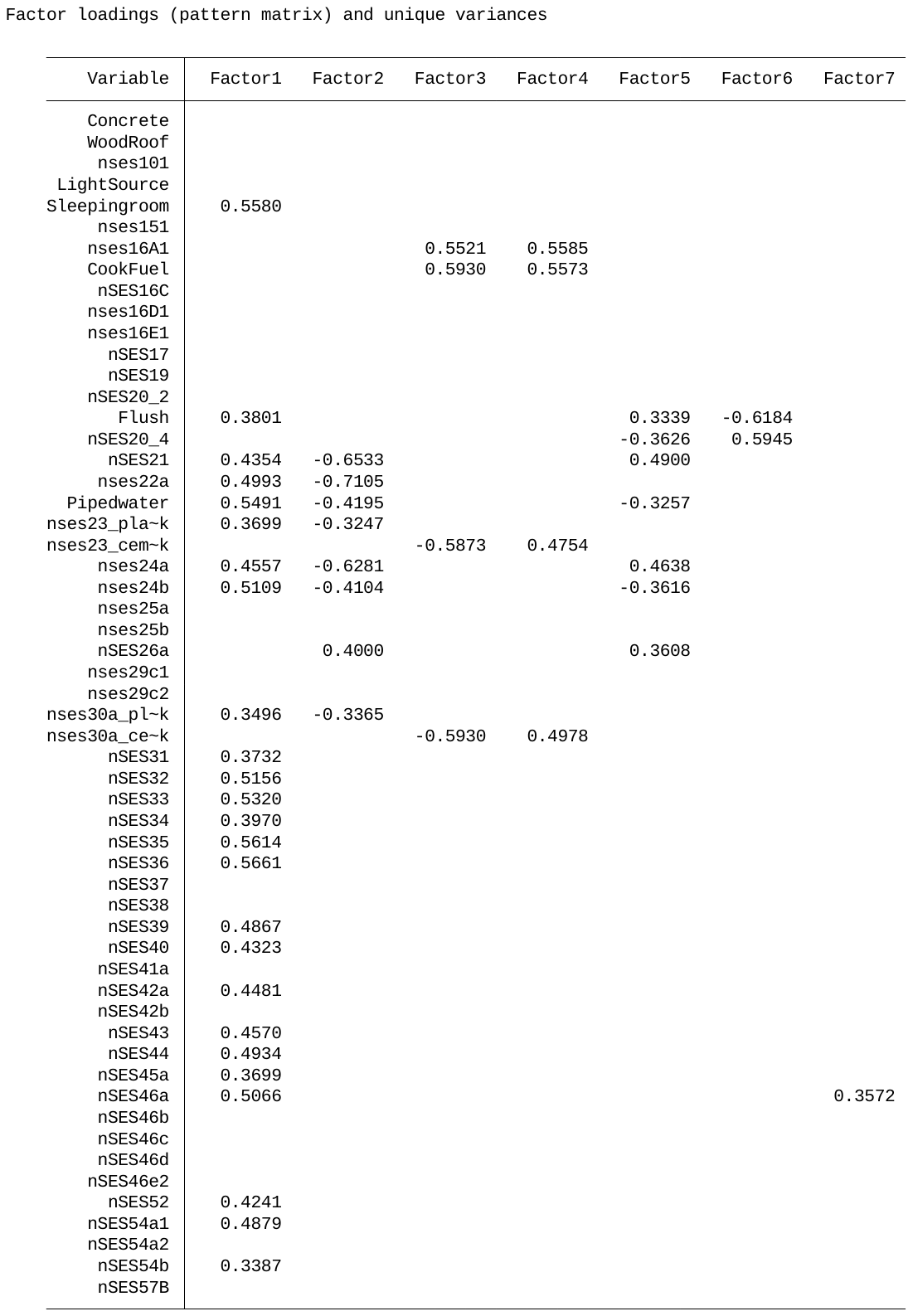

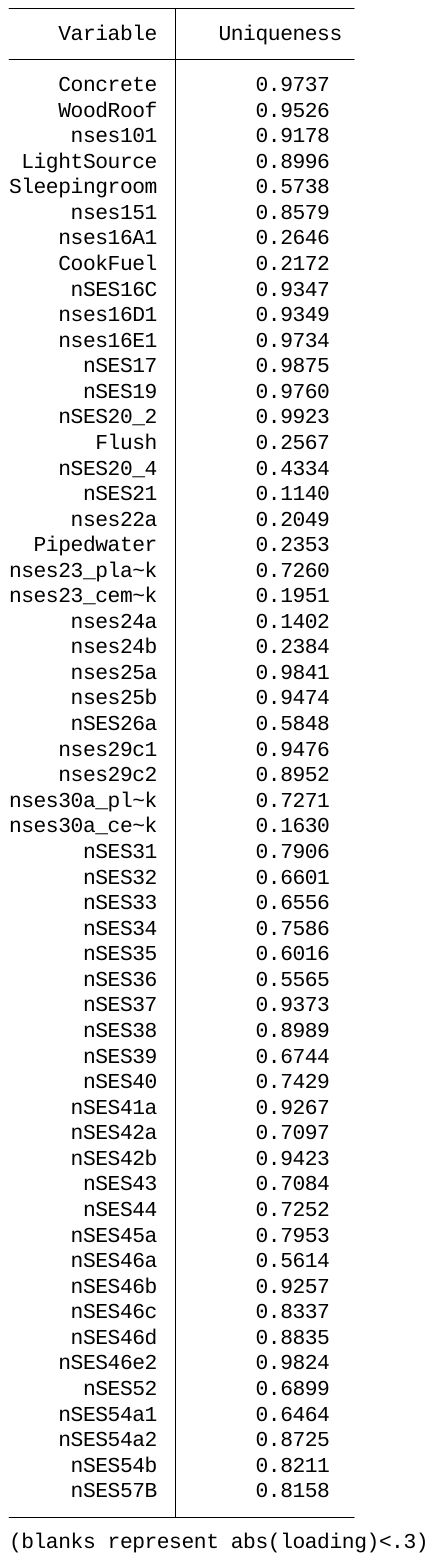

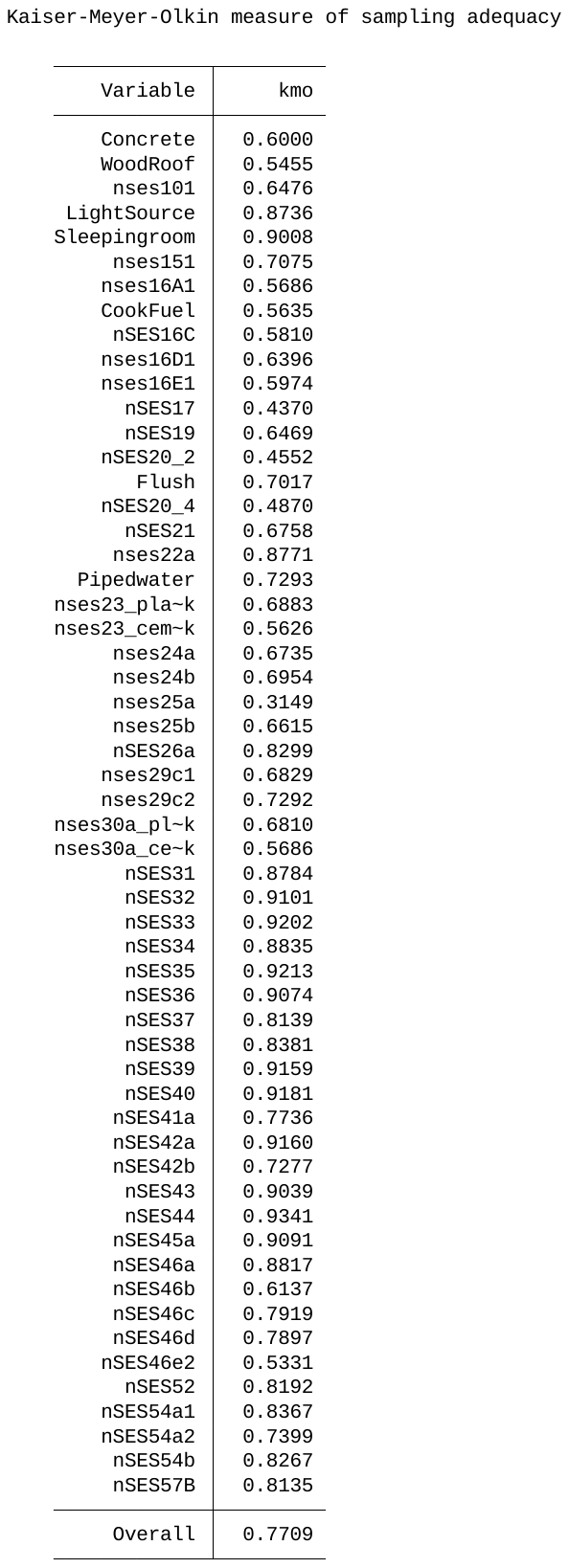
