## Supplemental Appendix 1 for "Rapid changes in population socioeconomic status indicators are unevenly distributed in a rural Pakistani village"

**Socioeconomic and Environmental Survey**

|  |  | | | | | Code of Supervisor | | | | | | **02** | |  | | Code of Interviewer | | | | | | | | | | **01b** | | | | __ __ / __ __ __ / __ __ | | | | | | | | Date of Interview | | **01a** |
| --- | --- | --- | --- | --- | --- | --- | --- | --- | --- | --- | --- | --- | --- | --- | --- | --- | --- | --- | --- | --- | --- | --- | --- | --- | --- | --- | --- | --- | --- | --- | --- | --- | --- | --- | --- | --- | --- | --- | --- | --- |
|  |  | | | | | | | | | | | Name of Respondent | | | | **03b** | |  | | |  | | |  | | | | |  | |  |  | |  | |  | | Respondent’s ID | | **03a** |
|  | Mother = 01; Grandmother = 02; NA = 97 | | | | | | | | | | | | | | | | |  | | | | |  | | | | What is your relationship to the child enrolled in WSHHI? | | | | | | | | | | | | | **03c** |
|  | Wife = 01; Mother/Mother-in-law = 02; Daughter/Daughter-in-law = 03; Step-daughter/adopted daughter = 04; Granddaughter = 05; Sister/Sister-in-law = 06; Aunt = 07; Niece = 08; Cousin = 09; Other (specify) | | | | | | | | | | | | | | | | |  | | | | |  | | | | What is your relationship to the head of the household (primary respondent)? | | | | | | | | | | | | | **03d** |
|  | If sufficient information is not ascertained from the first respondent, then second respondent will have to be selected. | | | | | | | | | | | | | | | | | | | | | | | | | | | | | | | | | | | | | | | |
|  |  |  |  | |  | |  |  |  |  | Secondary Respondent’s ID: | | | | | | **04b** | | |  | | | | | | | | | | | | | | | Secondary Respondent Name: | | | | | **04a** |
|  |  | | | | | | | | | | | | | | | | | | | | | | | | | | Questions answered by Secondary Respondent (List) | | | | | | | | | | | | | **04c** |
|  | Mother = 01; Grandmother = 02; Aunt = 03; Sister = 04; Father = 05; Grandfather=06; Uncle=07; Brother=08; Cousin=09; NA=97; Other (specify) | | | | | | | | | | | | | | | | |  | | | | |  | | | | Secondary respondent’s relationship to the child enrolled. | | | | | | | | | | | | | **04d** |
|  | Wife = 01; Mother/Mother-in-law = 02; Daughter/Daughter-in-law = 03; Step-daughter/adopted daughter = 04; Granddaughter = 05; Sister = 06; Aunt = 07; Niece = 08; Cousin = 09; Father = 10; Son/Son-in-law = 11; Step-son/adopted son = 12; Grandson = 13; Brother = 14; Uncle = 15; Nephew = 16; Head of Household (Self) = 17; Other (specify) | | | | | | | | | | | | | | | | |  | | | | |  | | | | What is your relationship to the head of the household (secondary respondent)? | | | | | | | | | | | | | **04e** |
|  | **Education Questions** | | | | | | | | | | | | | | | | | | | | | | | | | | | | | | | | | | | | | | | |
|  | Primary (Prep-5) = 01; Middle (6-8) = 02; Matric (9-10) = 03; Intermediate (FA, B.Sc., I.Com) = 04; College/University Bachelors (BA, BSc, B.Ed., B Com, MBBS, B.Pharmacy) = 05; University Masters or Above (MA. MSc, MBA, MPhil, PhD, etc.) = 06; Professional Training and Other Diplomas/Certificates (LHW, PTC, CT, Montessori, etc.) = 07; Madrasa = 08; None = 09 | | | | | | | | | | | | | | | | |  | | | | |  | | | | What is the highest level of education achieved in your *household*? (Multiple responses possible) | | | | | | | | | | | | | **05** |
|  | Yes = 01; No = 02 | | | | | | | | | | | | | | | | |  | | | | |  | | | | Are you currently attending school or university? | | | | | | | | | | | | | **06** |
|  | **Housing Questions** | | | | | | | | | | | | | | | | | | | | | | | | | | | | | | | | | | | | | | | |
|  | Earth/sand = 01; Wood = 02; Cement/concrete = 03; Other (specify): | | | | | | | | | | | | | | | | |  | | | | |  | | | | Main material of the floor (Observation) | | | | | | | | | | | | | **07** |
|  | Yes = 01; No = 02 | | | | | | | | | | | | | | | | |  | | | | |  | | | | Is the floor carpeted? (Observation) | | | | | | | | | | | | | **08** |
|  | No roof=01; Metal=02; Wood=03; RCC=04; Tiles=05; Slate=06; Other | | | | | | | | | | | | | | | | |  | | | | |  | | | | Main material of the roof? (Observation) | | | | | | | | | | | | | **09** |
|  | No walls=01; Mud=02; Wood=03; Cement/concrete=04; Stone=05; Brick =06;Other | | | | | | | | | | | | | | | | |  | | | | |  | | | | Main material of the exterior walls? (Observation) | | | | | | | | | | | | | **10** |
|  | Number of rooms | | | | | | | | | | | | | | | | |  | | | | |  | | | | How many rooms are there in your house? | | | | | | | | | | | | | **11** |
|  | Number of rooms | | | | | | | | | | | | | | | | |  | | | | |  | | | | How many rooms are used for sleeping? | | | | | | | | | | | | | **12** |
|  | Number of people | | | | | | | | | | | | | | | | |  | | | | |  | | | | Max number of persons sleeping in one room? | | | | | | | | | | | | | **13** |
|  | Yes = 01; No = 02 | | | | | | | | | | | | | | | | |  | | | | |  | | | | Does your household have electricity? | | | | | | | | | | | | | **14a** |
|  | Hours in summer  Hours in winter | | | | | | | | | | | | | | | | |  | | | | |  | | | | If yes, for how many hours each day is electricity absent? | | | | | | | | | | | | | **14b** |
|  |  |  |  |  |  |  |  |  |  |  |  |  |  |  |  |  |  |  | | | | |  | | | |  |  |  |  |  |  |  |  |  |  |  |  |  | **14c** |
|  | Generator = 01; Gas = 02; UPS system = 03; Chargeable lights = 04; Candles = 05; Lantern = 06; Stove = 07; Open fire = 08; Nothing = 09; Other (specify)  **[If not stove or open fire (7 or 8), skip to Q15]** | | | | | | | | | | | | | | | | |  | | | | |  | | | | In the case of discontinued power supply, what source does this household usually use? *(Please list in order of priority)* | | | | | | | | | | | | | **14d** |
|  | Wood =01; Gas=02; Kerosene=03; Trash = 04; Dung = 05; Other (Specify) | | | | | | | | | | | | | | | | |  | | | | |  | | | | If use stove or open fire for lighting, what type of fuel is used? | | | | | | | | | | | | | **14e** |
|  | Wood =01; Gas=02; Kerosene=03; Trash = 04; Dung = 05; Electricity = 06; Other (Specify) | | | | | | | | | | | | | | | | |  | | | | |  | | | | What are the primary sources of fuel used for heating in your household? (List in order of usage) | | | | | | | | | | | | | **15** |
|  | Open fireplace = 01; Gas stove = 02; Kerosene stove = 03; Electric stove = 04; Local bukhari = 05; BACIP improved stove with pipe and water heater = 06; BACIP improved stove with pipe but no water heater = 07; Other (specify) | | | | | | | | | | | | | | | | |  | | | | |  | | | | What types of cooking stoves are used in your house? (List in order of usage) | | | | | | | | | | | | | **16a** |
|  | Wood =01; Gas=02; Kerosene=03; Trash = 04; Dung = 05; Electricity = 06; Other (Specify) | | | | | | | | | | | | | | | | |  | | | | |  | | | | What are the primary sources of fuel used for cooking on each stove? (List in order of responses in 16a) | | | | | | | | | | | | | **16b** |
|  | Metal = 01; None = 02; Other (specify) | | | | | | | | | | | | | | | | |  | | | | |  | | | | Does each stove have a chimney? (List in order of responses in 16a) | | | | | | | | | | | | | **16c** |
|  | Same room as living area = 01; Separate room as living area = 02; Room outside main house = 03; Semi-covered area outside = 04; Courtyard/open area outside = 05; Other (specify) | | | | | | | | | | | | | | | | |  | | | | |  | | | | Where is each stove located in summer? (List in order of responses in 16a) | | | | | | | | | | | | | **16d** |
|  | Same room as living area = 01; Separate room as living area = 02; Room outside main house = 03; Semi-covered area outside = 04; Courtyard/open area outside = 05; Other (specify) **[If courtyard/open area outside (5), skip to Q18a]** | | | | | | | | | | | | | | | | |  | | | | |  | | | | Where is each stove located in winter? (List in order of responses in 16a) | | | | | | | | | | | | | **16e** |
|  | BACIP roof hatch = 01; Open window in ceiling = 02; Chimney hole in ceiling = 03; Window in wall = 04; None = 05; Other (specify) | | | | | | | | | | | | | | | | |  | | | | |  | | | | What type of ventilation is in the kitchen? (Mark all that apply) | | | | | | | | | | | | | **17** |
|  | **If household has a BACIP stove, skip to Q19** | | | | | | | | | | | | | | | | | | | | | | | | | | | | | | | | | | | | | | | |
|  | Yes= 01; No= 02 **(If no, skip to Q19(** | | | | | | | | | | | | | | | | |  | | | | |  | | | | Have you heard of the BACIP stove? | | | | | | | | | | | | | **18a** |
|  | Too expensive = 01; Difficult to use = 02; Don’t want to try another stove = 03; Doesn’t cook well = 04; Doesn’t heat room well = 05; Other (specify) | | | | | | | | | | | | | | | | |  | | | | |  | | | | Why don’t you have a BACIP stove? (Mark all that apply) | | | | | | | | | | | | | **18b** |
|  | Yes = 01; No = 02 | | | | | | | | | | | | | | | | |  | | | | |  | | | | Was there any smoke in the house at the time of the interview? | | | | | | | | | | | | | **19** |
|  | **Water and Sanitation Questions** | | | | | | | | | | | | | | | | | | | | | | | | | | | | | | | | | | | | | | | |
|  | No facility/field=01; Pit latrine(old)/Chukan=02; Twin Pit latrine(new-composting WASIP)=03; Flush to septic tank=04; Flush to pit latrine=05; Other | | | | | | | | | | | | | | | | |  | | | | |  | | | | What kind of toilet facility do members of your household usually use? | | | | | | | | | | | | | **20** |
|  | Yes = 01; No = 02 **(If no, skip to Q22)** | | | | | | | | | | | | | | | | |  | | | | |  | | | | Do you have access to water from a filtration plant? | | | | | | | | | | | | | **21a** |
|  | WRI (Zohara) = 01; WRII (Dar) = 02 | | | | | | | | | | | | | | | | |  | | | | |  | | | | If yes, which filtration plant? | | | | | | | | | | | | | **21b** |
|  | Channel = 01; Water filtration plant WRI (Zohara) = 02; Water filtration Plant WRII (Dar) = 03; Other (specify) : __________ | | | | | | | | | | | | | | | | |  | | | | |  | | | | What is the main source of water members of your household use for household tasks (e.g. washing, cooking, and cleaning)? | | | | | | | | | | | | | **22a** |
|  | Piped to dwelling = 01; Piped to yard/plot = 02; Public tap/stand pipe = 03; Fetch from channel = 04; Other (specify): ________________ | | | | | | | | | | | | | | | | |  | | | | |  | | | | How is this water delivered to your home? | | | | | | | | | | | | | **22b** |
|  | Matka (covered cooler) = 01; Uncovered cooler = 02; Gulk = 03; Covered bucket = 04; Uncovered bucket = 05; Jerry can = 06; Plastic bottle = 07; Other (specify): ____ | | | | | | | | | | | | | | | | |  | | | | |  | | | | Where do you store your water used for household tasks? (Multiple responses possible) | | | | | | | | | | | | | **23** |
|  | Channel = 01; Water filtration plant WRI (Zohara) = 02; Water filtration Plant WRII (Dar) = 03; Other (specify) : __________ | | | | | | | | | | | | | | | | |  | | | | |  | | | | What is the main source of drinking water for members of your household? | | | | | | | | | | | | | **24a** |
|  | Piped to dwelling = 01; Piped to yard/plot = 02; Public tap/stand pipe = 03; Fetch from channel = 04; Other (specify) ____________________ | | | | | | | | | | | | | | | | |  | | | | |  | | | | How is this water delivered to your home? | | | | | | | | | | | | | **24b** |
|  | In own dwelling = 01; In own yard/plot = 02; Within 1 km = 03; Within 2 km = 04; Beyond 2 km = 05 | | | | | | | | | | | | | | | | |  | | | | |  | | | | Where is your main drinking water source located? | | | | | | | | | | | | | **24c** |
|  | Woman = 01; Man = 02; Girl under age 15 = 03; Boy under age 15 =04 | | | | | | | | | | | | | | | | |  | | | | |  | | | | Main person fetching water from this source? | | | | | | | | | | | | | **24d** |
|  | : Hours  : Minutes  (If source is located on the premises, response is 97. If no response, code 98 for both( | | | | | | | | | | | | | | | | |  | | | | |  | | | | How long does it take to go there, get water, and come back in one trip? | | | | | | | | | | | | | **24e** |
|  | Yes = 01; No = 02 | | | | | | | | | | | | | | | | |  | | | | |  | | | | Do you consider this main source of drinking water to be safe for drinking without treatment? | | | | | | | | | | | | | **24f** |
|  | Gulk=01; Channel=02; Water filtration plant=03; Bottled=04; Other | | | | | | | | | | | | | | | | |  | | | | |  | | | | Where do you get your drinking water from when the main source is not available? | | | | | | | | | | | | | **25a** |
|  | Piped to dwelling = 01; Piped to yard/plot = 02; Public tap/stand pipe = 03; Fetch from channel = 04; Other (specify): ____________________ | | | | | | | | | | | | | | | | |  | | | | |  | | | | How is this water delivered to your home? | | | | | | | | | | | | | **25b** |
|  | Continuous = 01; Sometimes interrupted = 02  **(If continuous, skip to Q27)** | | | | | | | | | | | | | | | | |  | | | | |  | | | | If you receive piped water, is it continuous or interrupted? | | | | | | | | | | | | | **26a** |
|  | Hours per day in the summer  Hours per day in the winter | | | | | | | | | | | | | | | | |  | | | | |  | | | | If interrupted, how long (in hours per day) is the interruption? | | | | | | | | | | | | | **26b** |
|  |  |  |  |  |  |  |  |  |  |  |  |  |  |  |  |  |  |  | | | | |  | | | |  |  |  |  |  |  |  |  |  |  |  |  |  | **26c** |
|  | Days per week in the summer  Days per week in the winter | | | | | | | | | | | | | | | | |  | | | | |  | | | | ا How many days per week is water interrupted for? | | | | | | | | | | | | | **26d** |
|  |  |  |  |  |  |  |  |  |  |  |  |  |  |  |  |  |  |  | | | | |  | | | |  |  |  |  |  |  |  |  |  |  |  |  |  | **26e** |
|  | Yes = 01; No = 02  **(If no, skip to Q28)** | | | | | | | | | | | | | | | | |  | | | | |  | | | | Do you or did you pay for water you receive in your home? | | | | | | | | | | | | | **27a** |
|  | _________________________ PKR (per month)  OR _________________________ PKR (total) | | | | | | | | | | | | | | | | |  | | | | | | | | | If yes, how much do you pay per month or did you pay initially for piped water? | | | | | | | | | | | | | **27b** |
|  | Yes = 01; No = 02 **(If no, skip to Q29(** | | | | | | | | | | | | | | | | |  | | | | |  | | | | Purchase drinking water from outside your home? | | | | | | | | | | | | | **28a** |
|  | _________________________ PKR | | | | | | | | | | | | | | | | |  | | | | | | | | | If 28a is yes, how much do you pay per month? | | | | | | | | | | | | | **28b** |
|  | Yes = 01; No = 02 **(If no, skip to Q30(** | | | | | | | | | | | | | | | | |  | | | | |  | | | | Treat your water to make it safer to drink? | | | | | | | | | | | | | **29a** |
|  | Primary = 01; Secondary = 02; Both = 03 | | | | | | | | | | | | | | | | |  | | | | |  | | | | Do you treat the primary or secondary source? | | | | | | | | | | | | | **29b** |
|  | Let it stand and settle = 01; Use water filter = 02; Strain through a cloth = 03; Add bleach or chlorine = 04; Boil = 05; Other (specify) | | | | | | | | | | | | | | | | |  | | | | |  | | | | Usual treatment to make water safer to drink? | | | | | | | | | | | | | **29c** |
|  | Minutes ____; Until it just begins to boil =55; No response = 98; Not boiled=97 | | | | | | | | | | | | | | | | |  | | | | |  | | | | If you boil water for drinking, for how long? | | | | | | | | | | | | | **29d** |
|  | All household members = 01; Children less than 5 years = 02; Children 5 -14 years = 03; Those who are ill = 04; Other (specify) _________ | | | | | | | | | | | | | | | | |  | | | | |  | | | | Who uses the treated water? (Multiple responses possible) | | | | | | | | | | | | | **29e** |
|  | Matka (covered cooler)=01; Uncovered cooler=02; Gulk=03; Covered bucket=04; Uncovered bucket=05; Jerry can=06; Plastic bottle=07; Other | | | | | | | | | | | | | | | | |  | | | | |  | | | | Where do you store your drinking water? | | | | | | | | | | | | | **30a** |
|  | Yes = 01; No = 02 | | | | | | | | | | | | | | | | |  | | | | |  | | | | Do livestock or other animals have access to wherever you store your drinking water? | | | | | | | | | | | | | **30b** |
|  | **Household Survey** | | | | | | | | | | | | | | | | | | | | | | | | | | | | | | | | | | | | | | | |
|  | Yes = 01; No = 02 | | | | | | | | | | | | | | | | |  | | | | |  | | | | Q31-45a read: "Does your household have a __?" | | | | | | | | | | | | |  |
|  | Yes = 01; No = 02 | | | | | | | | | | | | | | | | |  | | | | |  | | | | iron | | | | | | | | | | | | | **31** |
|  | Yes = 01; No = 02 | | | | | | | | | | | | | | | | |  | | | | |  | | | | bed | | | | | | | | | | | | | **32** |
|  | Yes = 01; No = 02 | | | | | | | | | | | | | | | | |  | | | | |  | | | | chair | | | | | | | | | | | | | **33** |
|  | Yes = 01; No = 02 | | | | | | | | | | | | | | | | |  | | | | |  | | | | sofa | | | | | | | | | | | | | **34** |
|  | Yes = 01; No = 02 | | | | | | | | | | | | | | | | |  | | | | |  | | | | cupboard | | | | | | | | | | | | | **35** |
|  | Yes = 01; No = 02 | | | | | | | | | | | | | | | | |  | | | | |  | | | | table | | | | | | | | | | | | | **36** |
|  | Yes = 01; No = 02 | | | | | | | | | | | | | | | | |  | | | | |  | | | | electric fan | | | | | | | | | | | | | **37** |
|  | Yes = 01; No = 02 | | | | | | | | | | | | | | | | |  | | | | |  | | | | radio/transistor | | | | | | | | | | | | | **38** |
|  | Yes = 01; No = 02 | | | | | | | | | | | | | | | | |  | | | | |  | | | | computer | | | | | | | | | | | | | **39** |
|  | Yes = 01; No = 02 | | | | | | | | | | | | | | | | |  | | | | |  | | | | television | | | | | | | | | | | | | **40** |
|  | Yes = 01; No = 02 | | | | | | | | | | | | | | | | |  | | | | |  | | | | mobile phone | | | | | | | | | | | | | **41a** |
|  | Number of cell phones | | | | | | | | | | | | | | | | |  | | | | |  | | | | How many mobile telephones? | | | | | | | | | | | | | **41b** |
|  | Yes = 01; No = 02 | | | | | | | | | | | | | | | | |  | | | | |  | | | | refrigerator | | | | | | | | | | | | | **42a** |
|  | **If any one of Questions 42a and 42b are ‘1(Yes)’ skip to Q43** | | | | | | | | | | | | | | | | |  | | | | |  | | | | freezer separate from refrigerator | | | | | | | | | | | | | **42b** |
|  | In neighbor’s refrigerator (01); Corner of channel/ inside the Gulk (02); no food left (03); Throw remaining food in garbage (04); Other(specify) | | | | | | | | | | | | | | | | |  | | | | |  | | | | How do you store your cooked food? (Mark all that apply) | | | | | | | | | | | | | **42c** |
|  | Yes = 01; No = 02 | | | | | | | | | | | | | | | | |  | | | | |  | | | | watch/clock | | | | | | | | | | | | | **43** |
|  | Yes = 01; No = 02 | | | | | | | | | | | | | | | | |  | | | | |  | | | | Bank Account | | | | | | | | | | | | | **44** |
|  | **If no, skip to Q46** Yes = 01; No = 02 | | | | | | | | | | | | | | | | |  | | | | |  | | | | any motorized vehicles) | | | | | | | | | | | | | **45a** |
|  | Car = 01; Van (e.g. hi-top) = 02; Jeep = 03; Large commercial truck = 04; Small commercial truck/pick-up = 05; Large commercial bus = 06; Tractor = 07; Motorcycle = 08; Bicycle = 09; Other (specify) | | | | | | | | | | | | | | | | |  | | | | |  | | | | If yes, which types of vehicles? (Mark all that apply) | | | | | | | | | | | | | **45b** |
|  | :Cows  :Goats  :Sheep  :Chickens  ___________________________ :Other (specify)  **(Code 97 if none; code 98 if no response)** | | | | | | | | | | | | | | | | |  | | | | |  | | | | How many cows, goats, sheep, chickens or other animals does this household own? | | | | | | | | | | | | | **46** |
|  | **Medical Questions** | | | | | | | | | | | | | | | | | | | | | | | | | | | | | | | | | | | | | | | |
|  | Yes= 01; No= 02 **(If no, skip to Q48(** | | | | | | | | | | | | | | | | |  | | | | |  | | | | Have you or any of your family members suffered from any acute or chronic illness in the last month? | | | | | | | | | | | | | **47a** |
|  | _________________________ PKR | | | | | | | | | | | | | | | | |  | | | | | | | | | What was cost of medical treatment in past month? | | | | | | | | | | | | | **47b** |
|  | Bank loan=01; Loan from family/friends=02; Loan from WO/VO/LSO =03; Stopped treatment=04; Savings=05; Sold land=06; Other (specify) | | | | | | | | | | | | | | | | |  | | | | |  | | | | How did you arrange the amount? (Mark all that apply) | | | | | | | | | | | | | **47c** |
|  | Yes = 01; No = 02 **(If no, skip to Q49)** | | | | | | | | | | | | | | | | |  | | | | |  | | | | Have any illnesses reduced the working productivity of an earning family member (e.g. less hours worked/salary earned, private business or farming practice suffered) | | | | | | | | | | | | | **48a** |
|  | Hours per week | | | | | | | | | | | | | | | | |  | | | | |  | | | | If yes, how many hours per week are/were lost? | | | | | | | | | | | | | **48b** |
|  | Car = 01; Van (e.g. hi-top) = 02; Jeep = 03; Large commercial truck = 04; Small commercial truck/pick-up = 05; Large commercial bus = 06; Tractor = 07; Motorcycle = 08; Bicycle = 09; Walk = 10; Other (specify) | | | | | | | | | | | | | | | | |  | | | | |  | | | | In the case of a medical emergency, how would you normally travel to receive medical treatment? | | | | | | | | | | | | | **49** |
|  | Wife = 01; Mother/Mother-in-law = 02; Daughter/Daughter-in-law = 03; Step-daughter/adopted daughter = 04; Granddaughter = 05; Sister = 06; Aunt = 07; Niece = 08; Cousin = 09; Father = 10; Son/Son-in-law = 11; Step-son/adopted son = 12; Grandson = 13; Brother = 14; Uncle = 15; Nephew = 16; Head of Household = 17; Other (specify) | | | | | | | | | | | | | | | | |  | | | | |  | | | | In the case of medical emergency, who must make final medical decisions in the household? (In relation to the head of household. Multiple responses possible) | | | | | | | | | | | | | **50** |
|  | Yes = 01; No = 02; No **(If no, skip to Q52)**  **(No Answer = 98; Don’t Know = 99)** | | | | | | | | | | | | | | | | |  | | | | |  | | | | Do any members of your household smoke? | | | | | | | | | | | | | **51a** |
|  | :(Number of members)  **(No Answer = 98; Don’t Know = 99)** | | | | | | | | | | | | | | | | |  | | | | |  | | | | If yes, how many members smoke? | | | | | | | | | | | | | **51b** |
|  | Yes = 01; No = 02; **No Answer = 98; Don’t Know = 99** | | | | | | | | | | | | | | | | |  | | | | |  | | | | Do they smoke inside the house? | | | | | | | | | | | | | **51c** |
|  | Cigarette= 01; Cigarette with naswar = 02; Hookah = 03; Other(specify) | | | | | | | | | | | | | | | | |  | | | | |  | | | | What is smoked? (Mark all that apply) | | | | | | | | | | | | | **51d** |
|  | **Household Background** | | | | | | | | | | | | | | | | | | | | | | | | | | | | | | | | | | | | | | | |
|  | Number of members | | | | | | | | | | | | | | | | |  | | | | |  | | | | How many household members earn financially? | | | | | | | | | | | | | **52** |
|  | **Please fill out the table on the following page according to the corresponding questions and codes below:**   1) Which members of the household are generally earning members? (Even if unemployed right now. In relation to head of household.)   Wife = 01; Mother/Mother-in-law = 02; Daughter/Daughter-in-law = 03; Step-daughter/adopted daughter = 04; Granddaughter = 05; Sister = 06; Aunt = 07; Niece = 08; Cousin = 09; Father = 10; Son/Son-in-law = 11; Step-son/adopted son = 12; Grandson = 13; Brother = 14; Uncle = 15; Nephew = 16; Head of Household = 17; Other (specify)   2) What occupation does this member of your household currently hold? (Multiple responses possible) 3) How much does each individual earn in one month? 4) How many hours per week does each individual work? 5) What is this household member’s current employment status?)   Currently employed = 01; Underemployed (employed but seeking more work) = 02; Unemployed (usually earning, now out of work; with a desire and ability to work, so is seeking work and available for work) = 03; Unable to work (usually earning, now out of work; some impediment preventing her from working, so not seeking work) = 04   6) In what city does each earning member work?   Oshikhandass = 01; Gilgit = 02; Hunza = 03; Punjab = 04; Sindh =05; Balochistan =06; KPK =07; FATA =08 Other (specify)   7) If looking for more work, how many additional hours does he/she desire? 8) If unable to work, what is preventing him/her from working?   Not enough time for work = 01; Does not want to work = 02; Disability (specify) = 03; Illness (specify) = 04; Family trouble (specify, if desire to do so) = 05; Unable to find a job despite trying, so stopped looking = 06; Other (specify) | | | | | | | | | | | | | | | | | | | | | | | | | | | | | | | | | | | | | | | **53** |
|  | | | | **1** | | | | | | | **2** | | | | **3** | | | | **4** | | | | | | | | | **5** | | | | | **6** | | | | **7** | | **8** | |
| **Family Members** | | | | **Members** | | | | | | | **Occupation** | | | | **Monthly Salary** | | | | **Weekly Hours** | | | | | | | | | **Employment Status** | | | | | **City of Work** | | | | **Additional Hours Wanted** | | **Unable to Work** | |
| **No. 1** | | | |  | | | | | | |  | | | |  | | | |  | | | | | | | | |  | | | | |  | | | |  | |  | |
| **No. 2** | | | |  | | | | | | |  | | | |  | | | |  | | | | | | | | |  | | | | |  | | | |  | |  | |
| **No. 3** | | | |  | | | | | | |  | | | |  | | | |  | | | | | | | | |  | | | | |  | | | |  | |  | |
| **No. 4** | | | |  | | | | | | |  | | | |  | | | |  | | | | | | | | |  | | | | |  | | | |  | |  | |
| **No. 5** | | | |  | | | | | | |  | | | |  | | | |  | | | | | | | | |  | | | | |  | | | |  | |  | |
| **No. 6** | | | |  | | | | | | |  | | | |  | | | |  | | | | | | | | |  | | | | |  | | | |  | |  | |
| **No. 7** | | | |  | | | | | | |  | | | |  | | | |  | | | | | | | | |  | | | | |  | | | |  | |  | |
| **No. 8** | | | |  | | | | | | |  | | | |  | | | |  | | | | | | | | |  | | | | |  | | | |  | |  | |
| **No. 9** | | | |  | | | | | | |  | | | |  | | | |  | | | | | | | | |  | | | | |  | | | |  | |  | |
| **No. 10** | | | |  | | | | | | |  | | | |  | | | |  | | | | | | | | |  | | | | |  | | | |  | |  | |
|  | Annual agricultural Income | | | | | | | | | | | | __________________  PKR | | | | |  | | | | | | | What is the total monthly income of the household? | | | | | | | | | | | | | | | **54a** |
|  | ______________________________ PKR | | | | | | | | | | | | | | | | |  | | | | | | | What is the total monthly savings of the household? | | | | | | | | | | | | | | | **54b** |
|  | Wife = 01; Mother/Mother-in-law = 02; Daughter/Daughter-in-law = 03; Step-daughter/adopted daughter = 04; Granddaughter = 05; Sister = 06; Aunt = 07; Niece = 08; Cousin = 09; Father = 10; Son/Son-in-law = 11; Step-son/adopted son = 12; Grandson = 13; Brother = 14; Uncle = 15; Nephew = 16; Head of Household = 17; Other (specify) | | | | | | | | | | | | | | | | |  | | | |  | | | Who makes financial decisions in the family? (In relation to the head of household. Multiple responses possible) | | | | | | | | | | | | | | | **54c** |
|  | Yes =01; No = 02 **(If no, skip to Q56)** | | | | | | | | | | | | | | | | |  | | | |  | | | Do you have any financial debt? | | | | | | | | | | | | | | | **55a** |
|  | ______________________________ PKR | | | | | | | | | | | | | | | | |  | | | | | | | If yes, how much? | | | | | | | | | | | | | | | **55b** |
|  | Own = 01; Rent = 02 **)If rent, skip to Question 56f)** | | | | | | | | | | | | | | | | |  | | | |  | | | Do you own or rent your home? | | | | | | | | | | | | | | | **56a** |
|  | Years | | | | | | | | | | | | | | | | |  | | | |  | | | For how many years have you owned? | | | | | | | | | | | | | | | **56b** |
|  | Yes = 01; No = 02 **(If no, skip to Q57 (** | | | | | | | | | | | | | | | | |  | | | |  | | | Did you take any loan for purchasing this house? | | | | | | | | | | | | | | | **56c** |
|  | _________________________ PKR | | | | | | | | | | | | | | | | |  | | | | | | | How much was the loan? | | | | | | | | | | | | | | | **56d** |
|  | Yes = 01; No = 02 **)If no, skip to Q57(** | | | | | | | | | | | | | | | | |  | | | |  | | | Are you still making payments for this loan? | | | | | | | | | | | | | | | **56e** |
|  | _________________________ PKR | | | | | | | | | | | | | | | | |  | | | | | | | If you are on rent or house purchasing loan is still remaining, how much is the payment per month? | | | | | | | | | | | | | | | **56f** |
|  | Yes =01; No = 02 **(If no, skip to Q58)** | | | | | | | | | | | | | | | | |  | | | |  | | | Does your family own any land? | | | | | | | | | | | | | | | **57a** |
|  | :Kanals  :Marhalas | | | | | | | | | | | | | | | | |  | | | |  | | | How much land does your family own? | | | | | | | | | | | | | | | **57b** |
|  | Oshikhandass = 1; Astore = 2; Hunza = 3.; Bagrot = 04; Other (specify) | | | | | | | | | | | | | | | | |  | | | |  | | | Where is the land that your family owns located? (Multiple responses possible) | | | | | | | | | | | | | | | **57c** |
|  | :Farming(01)  :Shop/business(02)  :Residential rental(03)  ______________________________:Other (specify)  **97 for not applicable:** | | | | | | | | | | | | | | | | |  | | | |  | | | How many kanals and marhalas are used for each of the following purposes? | | | | | | | | | | | | | | | **57d** |
|  | :Years  :Months | | | | | | | | | | | | | | | | |  | | | |  | | | How long has your family lived in Oshikhandass? | | | | | | | | | | | | | | | **58a** |
|  | Bought land in Oshikhandass = 01; Relatives already in Oshikhandass = 02; Business/job moved here = 03; To be nearer Gilgit = 04; Better educational opportunities = 05; Economic difficulties = 06; Safety concerns in previous area = 07 | | | | | | | | | | | | | | | | |  | | | |  | | | Why did your husband or his family initially migrate to Oshikhandass? (Multiple responses possible) | | | | | | | | | | | | | | | **58b** |
|  | Yes = 01; No = 02 | | | | | | | | | | | | | | | | |  | | | |  | | | Did you migrate to Oshikhandass because of marriage? | | | | | | | | | | | | | | | **58c** |
|  | Hunza = 01; Bagrot = 02; Astore = 03; Haramosh = 04; Yasin = 05; Skardu = 06; Para Chanar = 07; Other (specify) | | | | | | | | | | | | | | | | |  | | | |  | | | Where are your household’s ancestors from?  (More than one response possible) | | | | | | | | | | | | | | | **59a** |
|  | Brusheski = 01; Shina = 02; Urdu = 03; Balti = 04; Pashtu = 05; Other | | | | | | | | | | | | | | | | |  | | | |  | | | What languages do you speak in your household? (Rank in order of most commonly used) | | | | | | | | | | | | | | | **59b** |
