## Supplemental Appendix 2 for "Rapid changes in population socioeconomic status indicators are unevenly distributed in a rural Pakistani village"

Lady Health Worker:_________________	Field Supervisor:____________

Household Identification Number:_______________	Date of Interview: ___D/____M/____Y

**(Current Household Members & Family Medical History Form)**

| HH member Number | Household (HH) Member Name | Sex | Date of Birth DD/MMM/YY | Age (in years) | Age (in months) | Age (in days) | Died since Jul 2011* |
| --- | --- | --- | --- | --- | --- | --- | --- |

Sex: 1. Male; 2. Female

*Died since July 2011: 1. Alive; 2. Died (or if registered later, died in the year before registration)

**Information about Family Medical History**

Have any immediate family members ever been diagnosed, or suspected to have any of the following medical conditions? If so, please list family member(s) and relevant information. Please include deceased family members, if they have been diagnosed and list the age at death, followed by D (i.e. 77D)

| HH member Number | Household (HH) Member Name | Sex | Age (in years) | Age (in months) | Age (in days) | Alive/Died |
| --- | --- | --- | --- | --- | --- | --- |
| 1. Cancer | | | | | | |
| 2. Diabetes | | | | | | |
| 3. Hypertension | | | | | | |
| 4. Cardiac Diseases including Heart Attack | | | | | | |
| 5. Blood Disorders like Hemophilia | | | | | | |
| 6. Genetic Diseases like Down Syndrome | | | | | | |
| 7. Mental Disorders like Depression | | | | | | |
| 8. Birth Defects like Cleft Palate | | | | | | |

Comments/Remarks
