## Supplemental Appendix 3 for "Rapid changes in population socioeconomic status indicators are unevenly distributed in a rural Pakistani village"

Lady Health Worker:________________	Field Supervisor:__________________

Structure Number:_________________	Household Number:_______________

Mother Identification Number:_______________	Date of Interview: ___D/____M/____Y

**(Children less than 5 years Enrolment Form)**

| Serial Number | Question | Answer |
| --- | --- | --- |
| 01 | Name and ID of the 1^st^ Child  Name and ID of the 2^nd^ Child  Name and ID of the 3^rd^ Child  Name and ID of the 4^th^ Child  Name and ID of the 5^th^ Child | **Date of Birth of the child**  _____Day/______Month/_______Year  _____Day/______Month/_______Year  _____Day/______Month/_______Year  _____Day/______Month/_______Year  _____Day/______Month/_______Year |
| 02 | Mother’s Name |  |
| a | Mother’s Age | ________________ Years |
| 03 | Mother’s Highest Level of Education |  |
| 04 | Mother’s Occupation |  |
| 05 | Father’s Name |  |
| a | Father’s Age | ________________ Years |
| 06 | Father’s Highest Level of Education |  |
| 07 | Father’s Occupation |  |

**Mother/Father’s Highest Level of Education**

1. Illiterate;	2. Can read and write;

3. Primary/Class 1 to 5;	4. Middle/ Class 6 to 8;

5. Secondary/Matric/Class 9 to 10;	6. Intermediate/Class 11 to 12;

7. Graduate/Class 13 to 14;	8. Post Graduate/Class 15 to 16
